# Rural dengue dynamics: the interplay of climate, built environment, and agriculture in Costa Rica

**DOI:** 10.64898/2026.02.12.26346219

**Authors:** Caroline Glidden, Emma K. Southworth, Talya Shragai, Diana Rojas-Araya, Adriana Troyo, Luis Enrique Chaves-González, Rodrigo Marín, Isaac Vargas Roldán, Erin Mordecai

## Abstract

Dengue is one of the world’s highest-burden arboviral diseases. Although classically considered an urban disease, many regions experience a substantial dengue burden in rural areas. The combined influence of long-term climate, short-term weather variation, local built environments, and land-use gradients on dengue dynamics in rural settings remains poorly understood, limiting our ability to predict shifting risk under global change. Here, we investigate these dynamics in Costa Rica to disentangle how these interacting socio-environmental factors shape rural dengue transmission. We first use 22 years of canton-level (admin-2) case data to establish that both dengue cases and incidence are consistently higher in rural than in urban districts. Then, using ten years of district-level (admin-3) monthly case data and a Bayesian hierarchical modeling framework, we identify the climatic and land-use features most strongly associated with dengue risk. Temperature underlies broad spatial patterns in dengue’s urban–rural distribution, while precipitation effects differ between coasts, reflecting intercoastal climate zone contrasts rather than interactions between urbanization and water availability. Given suitable climate, even modest levels of built infrastructure substantially increase risk, but the relationship plateaus at higher levels of building volume. Dengue risk is also elevated in areas with high agricultural crop cover at low and mid elevations but not at higher, cooler elevations. Together these results suggest that high risk of rural dengue in Costa Rica result from climate suitability aligning with baseline levels of built infrastructure, with agriculture potentially emerging as a distinct driver of rural dengue transmission.

## Introduction

Dengue is one of the most widespread and rapidly increasing arboviral diseases globally, causing an estimated 390 million infections each year (Bhatt et al., 2013). Dengue is canonically considered an urban disease; however, dengue transmission also occurs in rural areas, where it can exceed the case numbers and incidence in urban areas (Doum et al., 2020; Man et al., 2023; Saleh et al., 2020; Yoon et al., 2012). Rural transmission cycles are often overlooked in dengue research, leaving critical gaps in understanding how environmental, anthropogenic, and climatic factors shape rural transmission and how these patterns differ from urban areas.

Dengue virus (DENV) is primarily transmitted by the mosquito *Aedes aegypti*, but can also be transmitted by the globally invasive secondary vector *Aedes albopictus* (Garcia-Rejon et al., 2021; Ibáñez-Bernal et al., 1997; Kraemer et al., 2015; La Ruche et al., 2010). *Aedes* mosquitoes, and thus DENV exposure, are dependent on the environment. Temperature and humidity govern mosquito survival, reproduction, biting, and pathogen extrinsic incubation period (Brown et al., 2023; Mordecai et al., 2017, 2019). Breeding habitat availability is ultimately dictated by hydrological conditions and the presence of suitable containers: both mosquito species lay eggs and complete immature development in water-filled containers, primarily human-made containers such as tires, buckets, and plastic trash (Calderón Arguedas et al., 2012; Marín et al., 2009, 2013; Rojas-Araya et al., 2017), although *Ae. albopictus* and, less commonly, *Ae. aegypti* can breed in natural containers such as coconuts, leaf axils, bamboo, and tree holes and stumps (Egid et al., 2022; Pereira-dos-Santos et al., 2020).

Together, these ecological constraints influence the distribution of the two major dengue vectors and their capacity for viral transmission.

Differences in urban versus rural transmission patterns may arise from spatial correlations between climate and human settlement patterns. Temperature is one of the most consistently identified global drivers of dengue risk, with numerous studies documenting its influence on incidence and/or vector occurrence (Athni et al., 2024; Barboza et al., 2023; Caldwell et al., 2021; Childs et al., 2025; Damtew et al., 2023; Gibb et al., 2023; Hidalgo et al., 2026; Lowe et al., 2018; Mordecai et al., 2019). Many urban areas are concentrated in warm coastal lowlands or deltas and these areas are frequently epicenters of large dengue outbreaks (Bhatt et al., 2013; Gubler, 2011; The World Bank, 2010). Precipitation also drives transmission, but its effects can be nonlinear and spatially heterogeneous. For example, in typically drier regions, heavy rainfall or water storage during drought can increase risk by creating new larval habitats, whereas in wetter regions, excessive precipitation may reduce risk by washing away immature mosquito life stages (Cawiding et al., 2025; Harris et al., 2024; Moise et al., 2024; Morrow et al., 2010; Stewart Ibarra et al., 2013). As with temperature, spatial patterns of urban versus rural areas may be confounded with precipitation regimes, potentially producing climate-correlated rural–urban contrasts in dengue risk.

Alternatively, rural–urban differences may be caused by socio-ecological mechanisms unique to each landscape. Urban heat islands, for instance, can enhance mosquito development and DENV transmission up to an upper thermal threshold, after which conditions become unsuitable (Araujo et al., 2015; Sureshkumar & Shekhar, 2025; Wimberly et al., 2020), potentially producing contrasting effects of urban cover on dengue transmission depending on underlying temperatures. Tree cover can also have divergent effects: in urban settings, tree cover may provide thermal refuge and has been found to have higher larval occurrence (Troyo et al., 2009), whereas in rural landscapes, tree cover or forest proximity has been associated with reduced *Ae. aegypti* and *Ae. albopictus* and lower dengue risk (Farner et al., 2025; Gregory et al., 2022; Piaggio et al., 2024; Saager et al., 2023), potentially due to broader cooling effects or stronger biotic regulation by competitors and predators. Finally, socio-economic differences may interact with climate to produce local urban versus rural differences: access to piped water and other water infrastructure may diverge between urban and rural areas, and, consistent with recent dengue studies linking hydrological extremes, urbanization, and infrastructure (Li et al., 2023; Lowe et al., 2021), this may interact with precipitation to drive differences in urban versus rural transmission patterns.

Although less explored, agricultural landscapes may independently influence dengue risk through water-storage practices, limited sanitation infrastructure, vegetation structure, and human socioeconomics and behavior. Both *Ae. aegypti* and *Ae. albopictus* have been found in agricultural areas, where farming containers, discarded equipment, and crop plants (e.g., taro, pineapple) have been found to create vector habitats (Aryaprema, 2013; Calderón-Arguedas et al., 2015; Kadjo et al., 2025; Zahouli et al., 2017). Further, urban areas may have more reliable piped water and waste management than rural farmland, and as a result, agricultural communities may face greater exposure to vector habitats due to standing water storage. In many settings, rural populations engaged in seasonal agricultural work also experience high mobility, crowded or substandard housing, and limited access to healthcare and preventative tools such as insect repellent (Argaw et al., 2021; Bolaños et al., 2008; Morales-Gamboa, 2021). These differences could help explain why dengue incidence is higher in rural areas in some regions.

Costa Rica represents an important setting in which to investigate the drivers of rural DENV transmission. Dengue is endemic with a high burden in rural areas, and while *Ae. aegypti* has been established and contributed to transmission since at least 1993, *Ae. albopictus* has recently invaded and expanded throughout the country (Marín et al., 2009, 2012, 2013, 2014; Rojas-Araya et al., 2017). Costa Rica is a Central American country featuring a central mid–high elevation region—including the Central Valley and the Cordillera Central and Cordillera de Talamanca mountain ranges—surrounded by tropical lowlands with distinct precipitation patterns between its Caribbean and Pacific coasts. Urban centers concentrate in the cooler Central Valley highland (average elevation of 1,160 m.a.s.l.), whereas rural populations are distributed primarily across the warmer Atlantic and Pacific lowlands. These lowlands form a mosaic of forest, cropland, cattle pasture, and dispersed rural settlements, and include only three large urban centers (Liberia, Limón, and Puntarenas cities), which have high population densities but limited geographic extent. The alignment of climate gradients with rural settlement patterns and landscape heterogeneity provides an ideal opportunity to disentangle the respective environmental, climatic, and anthropogenic drivers shaping rural dengue risk.

Here, we use 22 years of country-wide annual canton-level (admin-level 2) data to describe the overlap between dengue cases and urban-rural landscapes. Zooming in, we then use ten years of country-wide monthly case data at the district level (admin-level 3) and a Bayesian spatio-temporal modeling framework to study potential determinants of the rural dengue spatial pattern in Costa Rica. Identifying the environmental characteristics of rural dengue, and whether they differ from urban settings, is crucial to understanding spatiotemporal changes in risk and informing targeted management strategies.

## Results

### Patterns in province-year dengue and urbanicity from 2000-2025

First, we evaluated broad, provincial trends in urbanicity and dengue incidence. From 2000-2025 (canton-level data), there were 421,966 reported cases of dengue in Costa Rica.

Throughout the time period where spatio-temporal modeling was applied to district-month cases (2011-2025, excluding 2012 and 2016), there were 185,877 total reported cases of dengue. We found that the rural provinces—those where populations were concentrated in smaller towns rather than large urban centers—had the highest total cases and incidence of dengue from 2000-2025 (Fig. 1). This pattern included the coastal provinces of Guanacaste, Puntarenas, and Limón. Provinces with a bimodal distribution in urbanicity, indicating about an equal portion of districts with both high urbanicity and high rurality, had the next highest case numbers, specifically San José and Alajuela provinces (Fig. 1). Alajuela consistently had the fourth-highest incidence over the past two decades, while San José’s incidence more closely resembled the lower-incidence provinces of Cartago and Heredia (Fig. 1). Further, the high-elevation province of Cartago followed a bimodal distribution in urbanicity, but had the lowest case numbers and incidence (Fig. 1), with the exception of a sharp increase in cases over the last five years: prior to 2021, average annual reported cases were 209, with 11 years (48% of annual time points) recording fewer than 100 cases; by contrast, average annual cases in 2021-2022 rose 8-fold to 1,927 (1,646 cases in 2021; 2,208 cases in 2022). This increase in cases was concentrated in Turrialba Canton, which spans an elevational gradient of 600-1,400 m.a.s.l. and is considered part of the Caribbean slope, bordering the Caribbean Limón province.

**Fig. 1.**
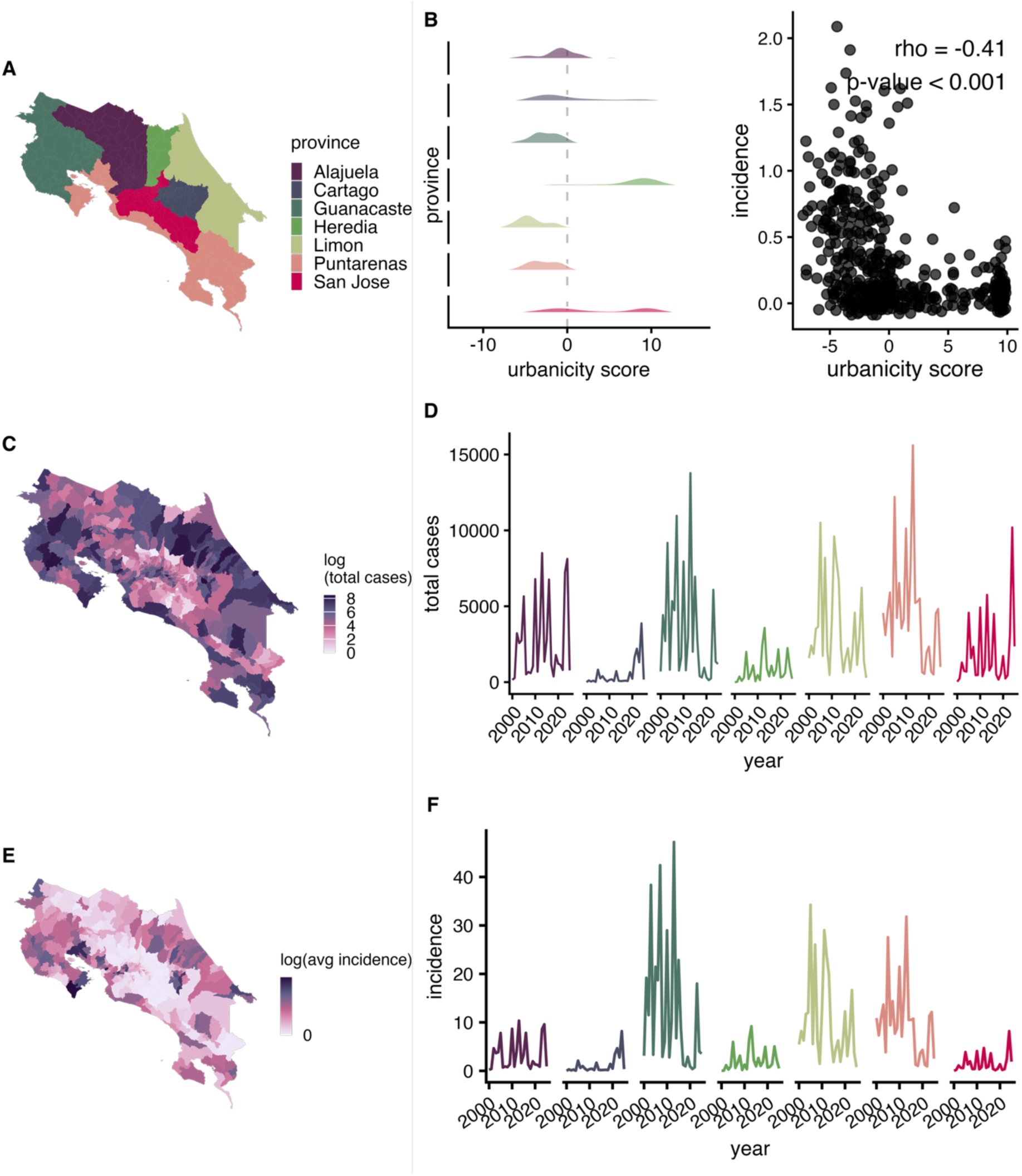
Spatio-temporal distribution of dengue and urbanicity. Map of provinces (A). Density plot of the average urbanicity score per district in each province (B), where low values indicate that people are concentrated in very rural areas, high values indicate concentration in urban centers, and 0 marks the threshold between rural and urban districts, and the relationship between average dengue incidence and average urbanicity score across districts. Total number of dengue cases per district from 2011–2025 (C) and per province-year from 2000–2025 (D). Average annual incidence (cases per 1,000 people) per district from 2011–2025 (E) and per province-year from 2000–2025 (F).

Heredia had the highest urbanicity and was among the provinces with the lowest incidence and case numbers; however, localized hotspots of dengue transmission were observed in northeastern regions, such as Las Horquetas and Puerto Viejo in the Sarapiquí canton, which also are considered part of the Caribbean slope. Supporting these results, we found a significant negative correlation between incidence and urbanicity at the canton scale (Fig. 1, ρ = -0.41, p-value < 0.001).

### Model Framework

Using a Bayesian spatio-temporal hierarchical model fitted with R-INLA (Lindgren & Rue, 2015), we quantified how climatic conditions (elevation, temperature, drought, and rainfall) and land use (built volume, crop cover, and forest cover) shape the spatial and temporal dynamics of district-month dengue risk in Costa Rica. Random effects were specified to absorb structured variation not captured by covariates: a cyclic seasonal term shared within provinces; and district-year level spatial effects specified as a BYM2 model, combining a spatially structured component (encoding the assumption that neighboring districts are more alike) with an unstructured component capturing district-specific heterogeneity.

Models were built sequentially—baseline random effects only, then climatic predictors, then land-cover predictors—with candidate models compared using DIC, WAIC, and the cross-validated logarithmic score, preferring a model that outperformed alternatives on at least two criteria. Temperature and precipitation were modeled as distributed lag non-linear effects (Gasparrini, 2014; Lowe et al., 2018). For elevation, drought, building volume, and forest cover, functional form (linear, logarithmic, or flexible non-linear) was selected identically. Because elevation was strongly collinear with both temperature (ρ = −0.85) and crop cover (ρ = −0.61), temperature was included as elevation-adjusted residuals and crop cover effects were estimated within elevation strata. Full model specification, priors, and selection procedures are detailed in the Methods.

### Statistical associations between district-month dengue risk and climate from 2011-2025

Temperature—both in its average relationship with elevation and in anomalies from it—improved model fit (Fig. 4; Supp Table 1). Dengue risk steeply decreased with elevation (Fig. 4; Supp Table 1). Similarly, after controlling for the temperature – altitude relationship, dengue relative risk increased nonlinearly with monthly minimum temperature anomalies at up to six months lag (Fig. 2; Supp Fig. 13; Supp Table 1).

**Fig. 2.**
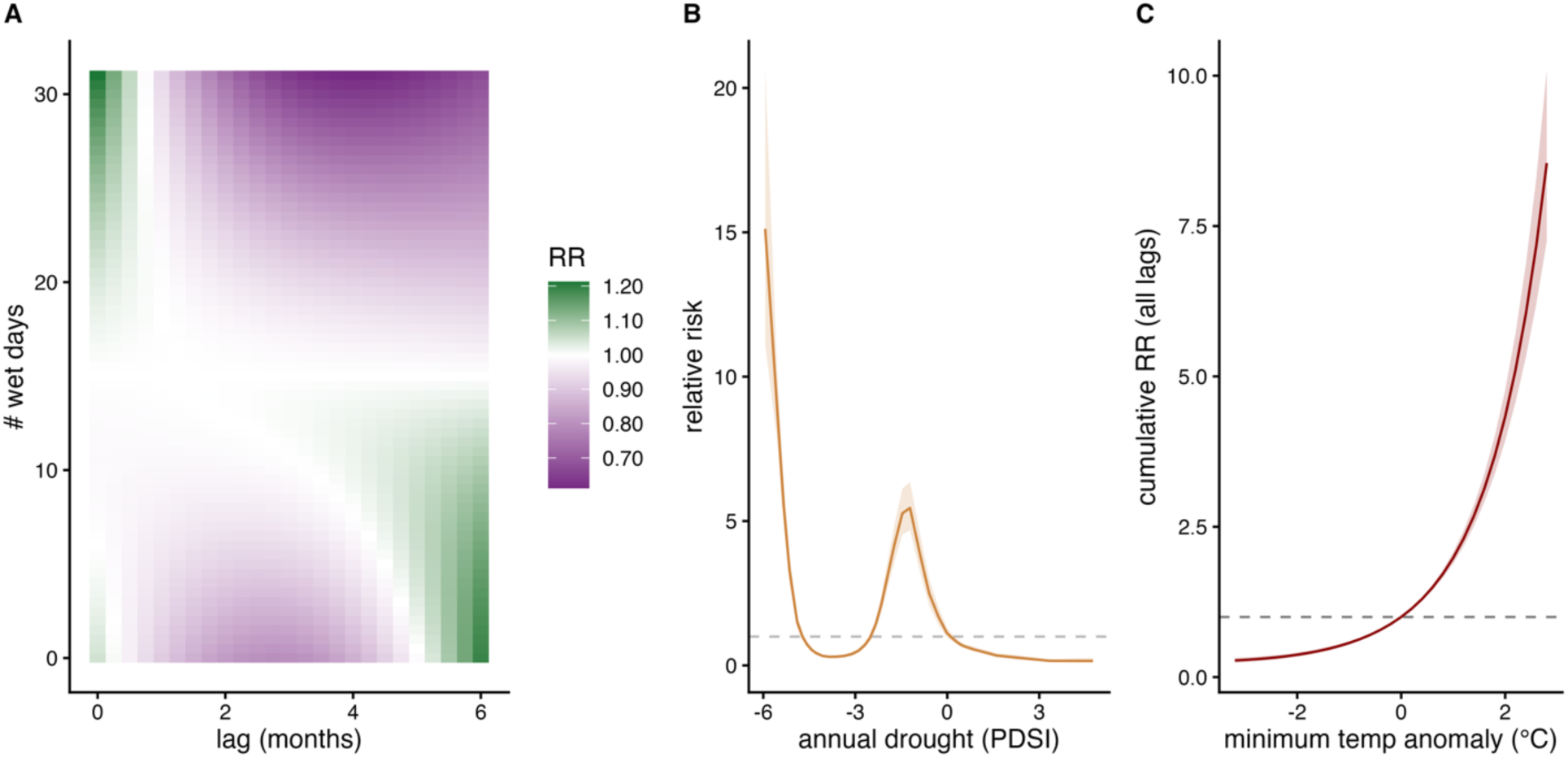
Lagged and non-linear effects of climate on dengue risk. Lagged non-linear effect of precipitation (number of wet days) (A), annual drought index (B), and monthly minimum temperature residual (°C deviation from elevation-based average minimum temperature) (C). Drought conditions are more severe when PDSI is < 0. Model selection was conducted to identify the number of lags of precipitation and temperature that best improved model fit. Allowing precipitation and temperature effects to lag up to six months improved model fit. Model covariates were included as center-scaled and relative risk is anchored by the mean of the covariate. The per-lag effect is shown for precipitation because its directional effect varies across lags, whereas the cumulative effect (across all six lags) is shown for temperature because it is directionally consistent across lags. Per-month and cumulative relative risk for each monthly climate variable are shown in Supp Fig. 13.

**Fig. 3.**
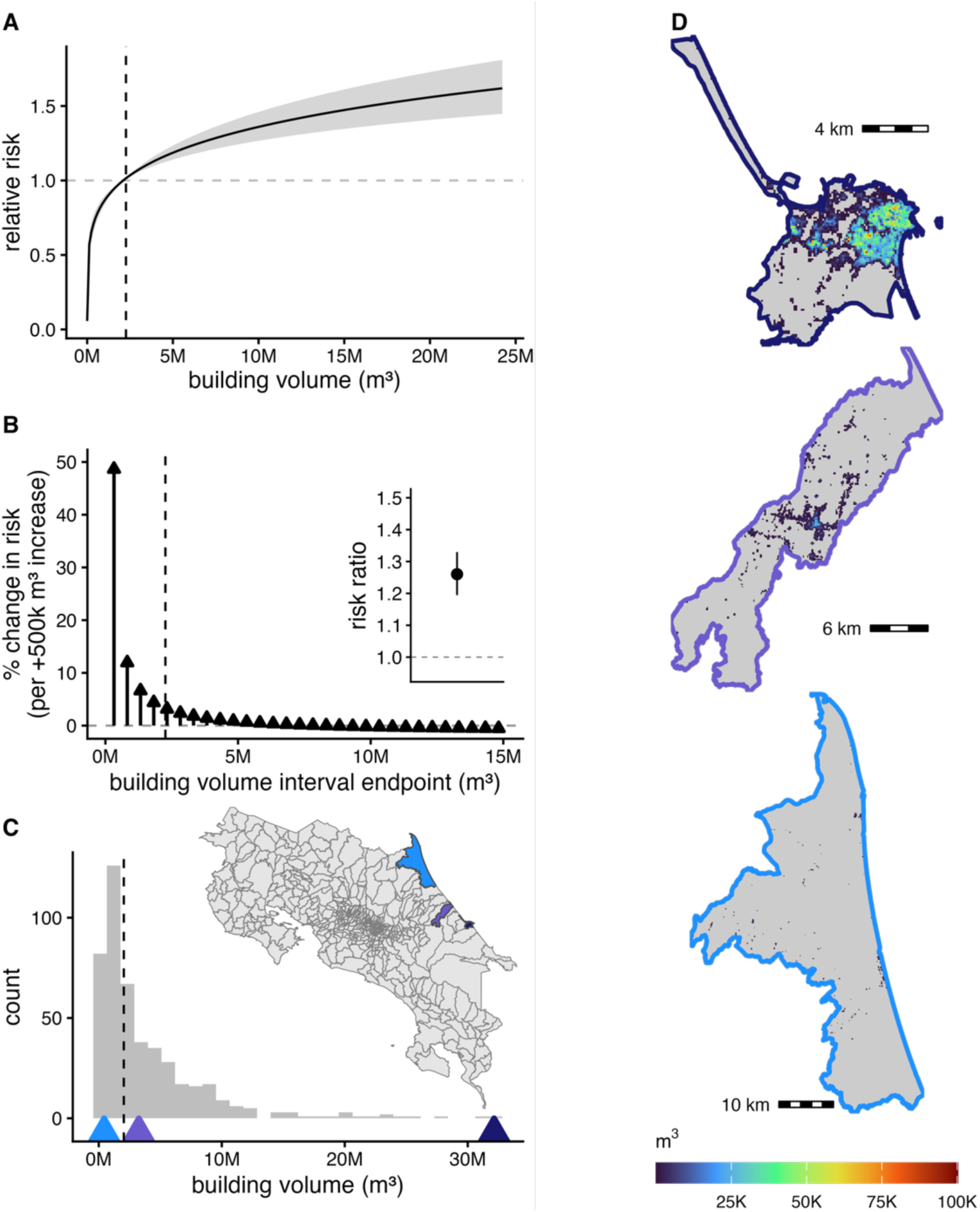
Effects of built environment on dengue risk. The relative risk of dengue as a function of mean building volume (A); the stepwise change in dengue risk with increasing building volume, expressed as the mean percent change in risk for each additional 500,000 m³ of building volume relative to the preceding interval (B, main panel), with the corresponding risk ratio for building volume shown as the model coefficient, which shows the rise in risk per standard deviation in building volume (B, inset); and the distribution of average building volume per district (C). Median building volume is indicated by vertical dashed black lines. Triangles along the base of the histogram indicate total district building volume for the three example districts shown in the maps to the right. Maps illustrate building volume at 100 m resolution in three coastal districts in Limón province (top, dark blue: Limón; middle, purple: Batán; bottom, light blue: Colorado). Median monthly dengue incidence (cases per 10,000 people) was 0.106 in Limón, 0.366 in Batán, and 0 in Colorado.

**Fig. 4.**
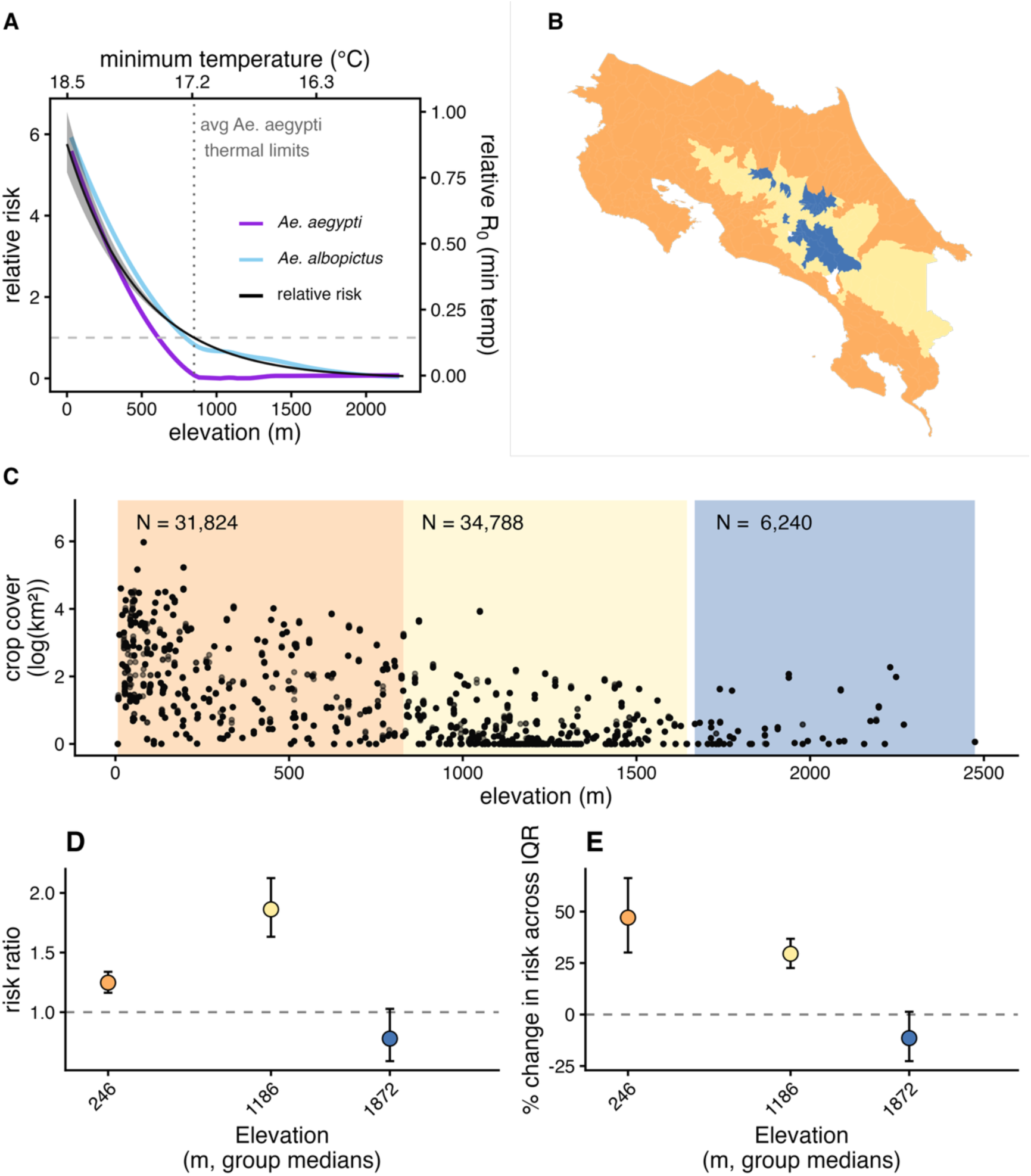
The effect of agriculture on dengue risk is highest at low and mid elevation. Relative risk of dengue by elevation (mean and 95% credible intervals, in black and gray, relative to the thermal suitability threshold for *Ae. aegypti*) is shown together with the temperature-dependent relative R₀ (Mordecai et al. 2017) by elevation (calculated for average minimum temperature along the elevation gradient) for *Ae. albopictus* (light blue) and *Ae. aegypti* (dark purple) at the mean minimum monthly temperature for a 20-year period by elevation (A). Panels also display elevation bins of average elevation per district distributed across Costa Rica (B), the co-linear relationship between elevation and agricultural land cover (C), the effect of agriculture on dengue incidence within each elevation stratum, shown as the change in risk associated with a 1 SD increase in log(crop cover) (D), and percent change in dengue risk associated with moving from the 25th to the 75th percentile of crop cover (i.e., a 1-IQR increase), estimated within each elevation stratum and illustrating the effect over the different ranges of crop cover within each stratum (E). Error bars in (D) and (E) represent 95% credible intervals around each estimate.

Precipitation and drought also predicted dengue incidence. We found that including the monthly number of wet days as a nonlinear model distributed over 6 months of lags and a measure of annual drought (average annual PDSI) improved model fit (Supp Table 2). Relative risk was highest with > 20 wet days 0-1 months before the observation and when there were few wet days 5-6 months before observation (Fig. 2; Supp Fig. 13). Dengue relative risk was highest during extreme and moderate drought years, decreasing during non-drought conditions (Fig. 2).

Drought, precipitation, and the combined temperature predictor (elevation as a long-term baseline and temperature anomalies capturing short-term deviations) improved out-of-sample dengue predictions in most districts, with strong spatial heterogeneity, whereas temperature anomalies alone (excluding elevation as a long-term baseline) improved predictions in slightly less than half of all districts (Fig. 5; Supp Table 6). The model-inferred mean drought-dengue relationship (i.e., the average association estimated across the posterior predictive distribution) decreased out-of-sample error, measured by a relative change in MAE by at least 1%, in 73% of districts, temperature (elevation + temperature anomalies) in 60% of districts, precipitation in 55% of districts, followed by temperature anomalies in 48% of districts (Supp Table 6). The inferred temperature-dengue relationship improved model performance at mid- to high elevations but reduced fit in lowland areas. In contrast, precipitation enhanced model performance in the western lowlands and reduced predictive performance along the eastern coast (Fig. 5).

**Fig. 5.**
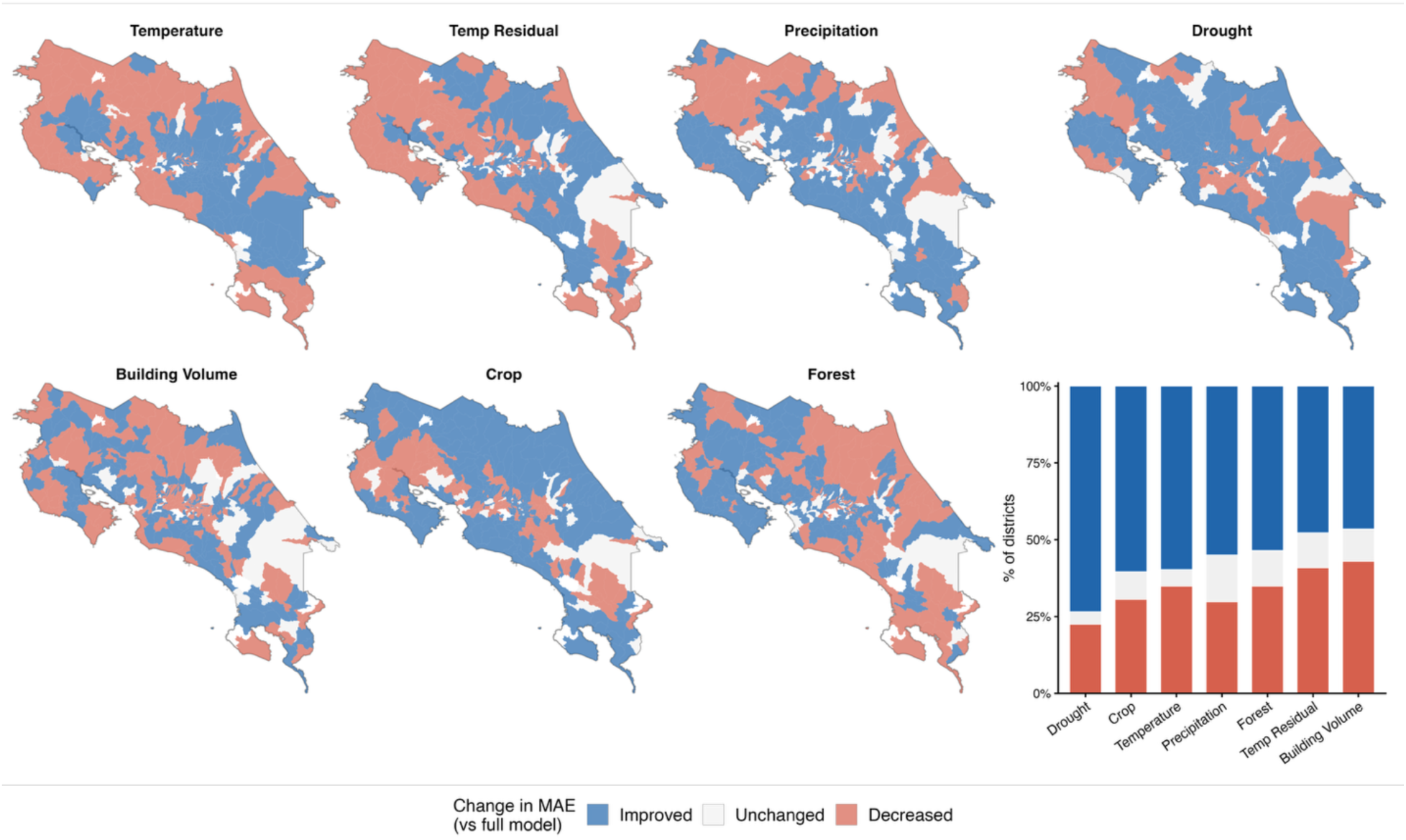
Spatial variation in variable importance, measured as the change in model error when omitting each focal variable. Maps indicate where inclusion of the variable changed relative MAE ((model_omitted variable_-model_full_)/model_full_) by at least 1%. Stacked bar graph indicates the proportion of districts where out-of-sample model predictions were improved, unaffected, or decreased. We define ‘Improved’ as relative MAE > 0.01, ‘Decreased’ as relative MAE < –0.01, and ‘No change’ as –0.01 ≤ relative MAE ≤ 0.01.

### Statistical associations between district-month dengue risk and land-use from 2011-2022

After accounting for climate, we found that including crop cover, forest cover, and building volume best improved prediction of dengue incidence; building volume improved model performance the most, followed by crop cover and then forest cover (Supp Table 3). Dengue risk increased nonlinearly with building volume. Specifically, building volume better predicted dengue risk as a log-transformed (logarithmic) term, compared to a linear term (Supp Table 3). The log transformation indicates that variation is best explained by a saturating relationship, where risk rises steeply at low-to-moderate building volumes before plateauing at higher levels (Fig. 3A). For example, dengue risk increases by 50% when comparing district–years with the minimum building volume (≈74,000 m³) to those with 500,000 m³ more building volume.

However, this effect sharply attenuates: once building volume reaches the mean level across the country (≈2,250,000 m³), further increases of 500 × 10³ m³ are associated with only a modest 0.41–4% rise in risk (Fig. 3B). This relationship implies that dengue transmission risk is elevated even when building volume is much lower than that observed in urban centers, consistent with the rural distribution of dengue cases (Fig. 3). Including building volume as a second-order random walk, which allows the data rather than an a priori transformation to determine the functional form, supported the robustness of the overall shape of the effect (Supp Fig. 15). The second-order random walk model additionally revealed uncertainty in the effect at very high building volume. Low-infrastructure districts were defined as those with limited access to electricity, sanitation services, or running water per the 2011 census. When these districts were excluded, dengue risk peaked at intermediate building volume rather than continuing to rise, and the upper tail was no longer significant (Supp Fig. 15). Excluding San José likewise rendered the effect at high building volume non-significant (Supp Fig. 15).

Crop cover generally had a positive association with dengue risk, where logarithmic crop cover best improved model fit, though the magnitude of this effect varied across elevation strata (Fig. 4D). The effect of agriculture was strongest in the mid-elevation stratum (∼750–1,750 m.a.s.l.), where a 1 SD increase in log(crop cover) was associated with an 86% higher risk of dengue (median RR = 1.86; 95% CI: 1.63–2.12). The effect was smaller but still positive in the low-elevation stratum (∼0–750 m.a.s.l.; RR = 1.25; 95% CI: 1.16–1.34), corresponding to a 25% higher risk per 1 SD increase. At high elevations (>1,750 m.a.s.l.), crop cover was not associated with dengue risk (RR = 0.77; 95% CI: 0.59–1.03). The range of crop cover was wider in low- than in mid-elevation districts (Fig. 4E). When comparing effects across the interquartile range (IQR) of crop cover, dengue risk in low-elevation areas increased by 47% (CI: 30%–66%) between the 25th and 75th percentiles of crop cover, corresponding to an increase from 1.2 to 20.4 km² (IQR = 19.2 km²). In contrast, in mid-elevation areas risk increased by 29% (CI: 22%–38%) across a much narrower span of crop cover, from 0 to 0.7 km² (IQR = 0.7 km²). In sum, agriculture had large effects on dengue risk at low to mid elevations even after controlling for the close association of crop cover to climatic variables, with the largest per-unit effects in mid elevations and the largest total effects in low elevations.

Forest cover best improved model fit as a flexible non-linear term, with dengue risk generally decreasing as forest cover increased. At very high (>50 hectares), data-sparse values of forest cover, however, the effect reversed direction, and credible intervals widened substantially. We found that the significant tail was driven mainly by districts in Cartago, Heredia, and Limón provinces (Supp Fig. 16). Removing all districts on the Caribbean slope (Limón province and the Turrialba and Sarapiquí cantons) reduced, but did not eliminate, the upturn at high forest cover, indicating that non-Caribbean-facing districts of Cartago and Heredia may also contribute to this association (Supp Fig. 16). Notably, the protective association at mid-ranges of forest cover depended on the inclusion of San José province, suggesting that the negative, potentially protective relationship is concentrated in San José (Supp Fig. 16). Overall, the effect of forest cover was spatially heterogeneous, indicating that the influence of forest cover on dengue risk likely depends on regionally specific interactions.

For land use, the model inferred crop ocover-dengue relationship contributed more consistently to improved district-level out-of-sample dengue predictions than building volume or forest cover (crop cover: 60% of districts; building volume: 46%; forest cover: 54%). Crop cover increased performance in the central part of the country and Caribbean coast but decreased performance in parts of Guanacaste, the province along the northwest Pacific coast (Fig. 5). Building volume improved model performance in districts with low volume but decreased performance in districts with high volume (Supp Fig. 16), consistent with the saturating relationship where relative risk flattens at moderate-high values of building volume and uncertainty in model predictions at high building volume (Fig. 5; Supp Fig. 16). Forest cover generally improved predictions in northwestern and central Costa Rica, but decreased predictions in the southwest and along the eastern coast. For districts with > 10k hectares of forest, where the final model indicates an increase in dengue risk, forest only increased model performance in four districts: Cahuita, Guácimo, and Valle La Estrella in Limón, and Pejibaye in Cartago.

Posterior predictive checks indicated that the model generally reproduced the overall distribution of dengue incidence, including the mean, variance, and the shape of the right-skewed tail (Supp Fig. 17). The 99th percentile of simulated counts closely matched the observed distribution, suggesting that the model captures the typical magnitude of high-incidence months (Supp Fig. 17). However, the model slightly underestimated the frequency of zero-incidence observations: the observed data contained 50,368 zeros, whereas posterior predictive simulations averaged ∼49,749 zeros (95% posterior range: 49,530–49,999).

## Discussion

Dengue risk was elevated in rural areas: provinces that had mostly rural populations experienced the highest dengue case numbers and incidence (Fig. 1). This pattern was associated with a combination of climatic and weather factors and nuanced land-use patterns. In lowlands, most of which are typically more rural, temperatures were generally conducive to transmission, whereas in urban areas, which are concentrated at higher elevations, long-term temperature and temperature variability more strongly determined dengue risk (Fig. 4A, Fig. 2, Fig. 5). A combination of long-term drought and short-term precipitation significantly increased dengue risk (Fig.3, Fig. 5). While dengue risk increased sharply with building volume (a proxy for the built environment) at very low levels, we found a low threshold above which dengue risk quickly saturated (Fig. 3B). Crop cover is associated with increased risk at low and mid elevations (0 m.a.s.l.-1,800 m.a.s.l.) but has no effect at high elevations (>1,800 m.a.s.l.) (Fig. 4C-D), and forest cover has regionally variable effects on dengue risk (Fig. 5). Taken together, this implies that the rural distribution of dengue burden in Costa Rica is driven by a combination of the climate suitability of lowland rural areas, the low amount of built volume needed to sustain transmission, and, potentially, the influence of agriculture.

Rural-urban patterns in dengue distribution are, at least in part, driven by spatial correlation between human settlement and temperature. Risk peaked at low elevations, declined with elevation, and then leveled off at higher altitudes (Fig. 4A), with anomalously high monthly minimum temperatures increasing dengue risk (i.e., deviations from the minimum temperature expected at that elevation; Fig. 2C). We also find that temperature improved out-of-sample predictions in mid to high elevation areas but not lowlands, suggesting that temperature more tightly constrains transmission in mid-high elevation areas (Fig. 5). Elevation and seasonal temperature are strong predictors of dengue and other *Ae. aegypti-*transmitted viruses globally (Gyawali et al., 2021; Watts et al., 2017), and have also been identified as the strongest correlates of dengue incidence across Costa Rica’s cantons (Mena et al., 2011). Consistent with this important influence of temperature, (Hidalgo et al., 2026) projected that dengue cases across Costa Rica will increase by mid-century (2035-2060) under the worst case (SSP5-8.5) climate warming scenario. Understanding temperature-imposed boundaries and how they may shift under climate change is critical for predicting timing and magnitude of dengue emergence into higher altitudes. In a supplementary analysis, we found that the median elevation of cumulative cases increased over time (Supp Fig. 14). Because elevation enters the model as a static, long-term climatology proxy, it does not track year-to-year change; however, the upward shift in the elevation of cases was instead captured by the annual spatial random effects, which became increasingly positively correlated with elevation over time (Supp Fig. 14). Consistent with an upward shift in suitable elevation, the final two outbreak years (2023–2024) showed higher altitudinal limits of thermal suitability for *Ae. albopictus* and *Ae. aegypti* than earlier years (Supp Fig. 14). Our results imply that the current climate-limited relative risk threshold for dengue in Costa Rica could extend with warming average temperatures, overlapping with large population centers such as San José.

Precipitation primarily shaped coastal differences but was not correlated with urban-rural settlement patterns. Relative risk of dengue was highest when there were many wet days in the current and previous month but decreased when there were dry conditions in the preceding two to six months (Fig. 2A), likely capture disease seasonality. Including precipitation improved out-of-sample performance within the central and Pacific cantons of Alajuela, Puntarenas, Guanacaste, and San José but decreased out-of-sample performance in the Caribbean province of Limón (Fig. 5), confirming that this wetter region has a distinct seasonality and response to precipitation than the rest of the country. These findings match (García et al., 2023), which found a significant correlation between precipitation and dengue in cantons in western Costa Rica, but no correlation between precipitation and dengue in the majority of the Caribbean cantons studied. Limón Province has a tropical monsoon climate that cycles between wet and wetter conditions, so abundant year-round rainfall provides ample vector habitat. Thus, other drivers likely dominate outbreak timing and magnitude in Limón. Temperature residuals — which capture fine-scale variation not explained by monthly climate averages — improved out-of-sample predictions in Limón but not in provinces with more pronounced dry and rainy seasons, such as Guanacaste on the Pacific coast (Romero-Vega et al., 2023), supporting this interpretation. Research in Puerto Rico and the Philippines similarly find the magnitude and direction of the effect of precipitation on dengue to be highly dependent on the underlying seasonality of the region (Cawiding et al., 2025). Overall, precipitation influences dengue dynamics in Costa Rica but does not explain the urban–rural gradient in incidence and burden during our study period.

Building volume had a positive but saturating effect on dengue risk, indicating that dengue can be sustained outside of urban areas. Dengue risk increased rapidly with initial increases in built volume and then plateaued, suggesting that DENV transmission in Costa Rica requires only a low threshold level of built infrastructure—such as that found in low to moderately urbanized areas like Batán, Costa Rica—to sustain transmission (Fig. 3). Including building volume improved out-of-sample performance only in districts with low to moderate built volume (Supp. Fig. 19), indicating that given adequate climate, dengue transmission depends on human-built environments but not necessarily highly urbanized settings. Even modest built environments may affect risk through altered container availability, increased temperatures via impervious surfaces, or modified human behavior (Wimberly et al., 2020). Supporting this finding, research within the city of Puntarenas, one of the three urban centers in the lowlands, found that *Ae. aegypti* was negatively correlated with built cover (Fuller et al., 2010), which may be due to fewer residential areas or improved sanitation in the city center. Troyo et al. (2009) also found abundant *Ae. aegypti* larvae in moderately built-up areas, particularly those with tree cover. On a regional scale, studies in the Peruvian Amazon have found that highway paving increased dengue cases in the Madre de Dios department (administrative level-2) despite having only one urban center (Singleton et al., 2026), suggesting only a baseline level of built infrastructure is needed for transmission, and that *Ae. aegypti* populations established in rural communities in the northern region of the Peruvian Amazon (Fikrig et al., 2025).

Our primary analysis lacked variables that could identify heterogeneity in dengue risk within built environments, such as water management practices (Arguedas et al., 2009; Gurevitz et al., 2021; Lowe et al., 2021), structural features of houses (Chastonay & Chastonay, 2022), and mobility (Shragai et al., 2022; Stoddard et al., 2013), as these variables are not available for the time period and spatial resolution of our study. However, a sensitivity analysis using census-derived infrastructure data (electricity, sanitation, and running water) suggested that the positive association between dengue risk and high building volume was driven largely by low-infrastructure districts: when these were excluded, risk peaked at intermediate building volume and the effect at high building volume was no longer significant (Supp Fig. 15). This suggests that infrastructure quality may modulate dengue risk within high building volume areas, though the use of outdated census data limits interpretation. Future work should integrate contemporaneous census and socioeconomic data with dengue case records to better characterize this relationship. Pinpointing heterogeneity would enable more targeted vector control and clarify whether transmission in rural built environments differs from urban built environments.

Crop cover was the only covariate in our analysis to suggest mechanisms that may be specific to rural areas, as the remaining urban-rural differences in dengue risk were largely explained by shared climatic and environmental drivers. Crop cover was strongly associated with increased dengue risk at low and mid elevations, even after accounting for the relationship between elevation and crop cover. Our correlational analysis cannot identify the mechanisms underlying this relationship, but several may be at play. Agricultural activity could reinforce socio-economic divides that affect services such as trash collection and housing quality; increase container availability through agriculture-related water management practices; and/or provide biotic breeding habitats, such as leaf axils, stumps, and coconut shells. Previous literature has found *Ae. aegypti* and *Ae. albopictus* in agriculture-based storage containers in farmlands and directly in crops (Aryaprema, 2013; Calderón-Arguedas et al., 2015; Kadjo et al., 2025) and existing literature shows that migrant workers in Costa Rica have limited access to social services and occupy more precarious housing (Bolaños et al., 2008; Voorend et al., 2023). In mid-elevation areas, agricultural activity could create warmer local microclimates (Senior et al., 2017) (Supp Fig. 18), making farms more suitable for dengue transmission than adjacent forested areas.

This mechanism may explain the sharper effect of crop cover at mid-elevation areas where climate suitability is more variable in space. While *Ae. aegypti* and *Ae. albopictus* have been previously observed in agricultural areas (Calderón-Arguedas et al., 2015; Kadjo et al., 2025; Zahouli et al., 2017), our study is among the first to identify a country-wide, multi-year association between crop cover and dengue risk. Further empirical work to uncover these mechanisms is important for identifying agricultural pathways of dengue transmission.

Consistent with the building volume sensitivity analysis, another important next step would be to incorporate district-level social vulnerability data. Socioeconomic inequality and public health infrastructure deficiencies act as risk multipliers through inadequate waste management, irregular water supply, and limited healthcare access (Alvarado-Prado et al., 2019), all conditions which are likely prevalent in agricultural areas (Bolaños et al., 2008; Voorend et al., 2023) and consistently highest in Costa Rica’s coastal provinces (MIDEPLAN, 2024). The human poverty index was already identified as a strong canton-level predictor of dengue incidence in Costa Rica from 1999–2007 (Mena et al., 2011), suggesting that socioeconomic conditions, rather than or in addition to crop cover directly, may underlie the rural dengue patterns we observe. Incorporating social vulnerability metrics would help to clarify whether the agriculture–dengue association reflects crop cover per se or underlying socioeconomic conditions, better differentiate risk in areas with mid-to-high built volume, and improve prediction at finer spatial scales.

Several limitations of these analyses should be considered. Dengue case detection is imperfect and under-reported, primarily relying on clinical diagnosis and epidemiological nexus (i.e., if there is an ongoing outbreak or there are other cases in the area). Other arboviruses that cause overlapping symptoms, such as Zika virus (ZIKV), chikungunya virus (CHIKV), and the recently detected Venezuelan Equine Encephalitis Virus (VEEV) may be misdiagnosed as dengue (Valles-Morera et al., 2025). This is particularly relevant given that Costa Rica experienced a major Zika outbreak in 2016, and asymptomatic or subclinical dengue infections may not have been captured by surveillance (Ministerio de Salud Costa Rica, 2023; Soto-Garita et al., 2024). Our study relies solely on case data and does not include entomological observations, meaning that we can only propose hypotheses linking epidemiological patterns to underlying vector mechanisms. These relationships should be further tested through targeted field studies and causal analyses to determine the relative contributions of *Ae. aegypti* and *Ae. albopictus* and their varying ecologies. Temporal data gaps may limit interpretation. District-level dengue data were unavailable for 2012 and 2016, and because our analysis spans only a decade, it captures relatively few large outbreak years. However, although we omitted 2016 due to missing data, this choice may have also reduced bias associated with misdiagnoses during the concurrent Zika outbreak. Our land-use variables were also non-specific, which could dilute relationships or mask mechanisms. For instance, our measure of forest cover broadly covers undisturbed moist tropical forest, and only some forest ecosystems may interact meaningfully with dengue dynamics, while our crop cover variable does not capture plantation agriculture, which may uniquely influence dengue dynamics and transmission.

## Conclusions

Costa Rica is an ideal setting for examining how long-term climate trends, short-term weather variability, and landscape features interact to drive dengue transmission in a country where the disease burden is largely rural. Environmental drivers of dengue are currently concentrated in rural areas, but this does not preclude the potential for dengue to emerge in urban settings. Like other tropical, dengue-endemic regions, this area is undergoing rapid global change as land use intensifies in agricultural areas, the climate warms, new vectors and virus serotypes circulate, and human populations grow and increase their mobility. In Costa Rica, the large urban populations living just above the elevational suitability limit for dengue raises a major risk for large outbreaks at higher elevations with warming minimum temperatures. The role of agriculture at the urban interface in these areas may serve as a point of emergence, expediting the increase in dengue cases in central regions. This pattern may be parallel other countries in the Americas, where large urban centers are currently out of the elevational reach of dengue but might become more suitable as climate warming progresses, such as in Mexico City, Mexico and Guatemala City, Guatemala (Childs et al., 2025). In this context, Costa Rica serves as an informative case for the Americas, where rural and peri-urban dengue transmission, often occurring in settings with limited surveillance and vector control capacity, remains an underappreciated contributor to regional disease burden under global change.

## Methods

### Dengue case data

Our dengue case data were provided by the Costa Rica Ministry of Health from their national surveillance system, which mandates that healthcare providers report probable and confirmed dengue cases. Probable dengue cases are diagnosed based on clinical symptoms and epidemiological context (e.g., if there is ongoing transmission in the area) (Ministerio de Salud Costa Rica, 2023). The Ministry of Health confirms a subset of cases at a national laboratory using molecular diagnostic tools on serum samples (e.g., PCR) (Ministerio de Salud Costa Rica, 2023). The Ministry of Health maintains a patient-level case report database that includes the district (admin-3) and date of diagnosis of each reported probable and confirmed case. We obtained deidentified case data for 2011-2025, but omitted 2012 and 2016 data as there were missing district identifiers (2012 data) and missing data in cantons where dengue had been known to occur (2016 data). Overall, the data include 72,852 district-months across ten years and 467 districts. Costa Rica currently has 492 districts; however, district boundaries were redrawn throughout the study period, often through subdivision into smaller districts. When feasible, we aggregated environmental and case data to align with the geographic boundaries of districts as defined at the start of the study. Some districts were omitted because it was not feasible to track their geographic boundaries over time or because they were located outside mainland Costa Rica. This study was approved by the Stanford University Institutional Review Board under protocol #85825 (approved March 31, 2026). The IRB granted waivers of informed consent and assent under 45 CFR 46.116(f)(3), given the minimal risk nature of the research.

In addition, we obtained weekly data at the canton level (admin-2) for the 2000-2020 period from Barboza et al. (2023). However, as the spatial resolution of cantons is too coarse to discern local land-use relationships, we only used these data to evaluate provincial trends over time and validate total number of cases per region-year in the district-level data. Visualizations of temporal and spatial trends in raw data are included in Supp. Figs. 1-2.

### Environmental data

We matched the district-level dengue case data with climate and land-use variables to assess their relationship with dengue risk. Climate variables included elevation (as a proxy for long-term average temperature), monthly minimum temperature, average annual Palmer Drought Severity Index (PDSI; a drought index representing climatic water balance), the number of wet days, which was calculated by summing the number of days in which total rainfall (mm) in the district-month exceeded the country median. Averages or sums were calculated in space across all pixels in the district. Monthly temperature variables (∼11 km resolution) were obtained from ERA5-Land (Muñoz Sabater, 2019), drought (measured as Palmer Drought Severity Index, PDSI, ∼4 km resolution) from TerraClimate (Abatzoglou et al. 2018), and precipitation (∼5 km resolution) from CHIRPS (Funk et al., 2015). Crop cover (annual, 300 m resolution) was downloaded from European Space Agency Climate Change Initiative Medium Resolution Land Cover data (Defourny, 2019). Crop cover included the categories rain-fed agriculture (code 10) and agricultural mosaic (>50% crop cover intermixed with other vegetation cover) (code 30).

Crop cover data only extended until 2022, as such, 2022 crop cover was used for 2023-2025. Crop cover only changed marginally over the study period (Supp Fig. 4), so using this approach likely closely represents crop cover in the final years of the study. Forest cover was downloaded from JRC Tropical Moist Forest v2025 dataset (∼30m resolution) (Vancustsem et al., 2020); only the undisturbed forest layer was used.

We obtained three different data layers from the Global Human Settlement Layer dataset (Pesaresi & Politis, 2023; Pesaresi et al., 2024; Pesaresi et al., 2024; Schiavina et al., 2023): building volume, population size, and degree of urbanization. For describing the built environment in our statistical analysis, we summed the total annual built volume per district-year (100m resolution). We used built volume as an explanatory variable in our model in lieu of the more traditionally used urban cover, as built volume captures the footprint and height of all buildings, including in low-density areas not considered urban. Using this variable allows us to draw inference on the threshold of the built environment needed for dengue transmission, even in more rural towns. We used the human population size per district-year for calculating dengue incidence and deriving the number of people within each urban class. Because GHSL data layers are available in 5-year intervals, we linearly interpolated the rasters across the study period to obtain a unique building volume and population count raster per year before aggregating data to the district level. Finally, we used the Degree of Urbanisation classification to describe the spatial distribution of the rural-urban gradient across Costa Rica. This layer classifies pixels as very low-density rural, low-density rural, rural cluster, suburban or peri-urban, semi-dense urban cluster, dense urban cluster, or urban center for the years 2000, 2005, 2010, 2015, 2020, and 2025, following UN Statistical Commission methodology based on population size, density, and built-up area. We selected this classification scheme because it is globally standardized, facilitating comparability and generalizability of our results. Visualizations of temporal and spatial trends in raw data are included in Supp. Figs. 3-11.

### Rural to urban gradient description

To qualitatively compare urbanicity with dengue cases and incidence over time we estimated the level of urbanization for every district from 2000 to 2025. Urbanicity was quantified based on how each district’s population was distributed across environments ranging from rural to urban, producing a continuous measure of exposure to different levels of urbanization. We applied Principal Component Analysis (PCA) to reduce these variables into a single continuous axis representing the urban–rural gradient. Scores along the first PCA axis were interpreted as district-level urbanicity scores, where lower values indicate populations concentrated in increasingly low-density rural areas, and higher values indicate populations concentrated in increasingly dense urban areas. Specifically, the negative scores correspond to districts where people are largely distributed across very low-density rural areas, values near zero represent semi-urban districts, and positive scores indicate districts where people are concentrated in urban centers. PCA was conducted using the *vegan* package in R v 4.3.2 (Oksanen et al., 2025), and all environmental data were processed and cleaned in Google Earth Engine using the Python API.

### Bayesian Spatio-temporal Hierarchical Model

#### Baseline model

Following Lowe et al. (Lowe et al., 2021) and Gibb et al. (Gibb et al., 2023), we fitted a spatio-temporal hierarchical model in a Bayesian framework using the *INLA* package in R v 4.3.2 (Lindgren & Rue, 2015; *R-INLA Project*, n.d.). This approach offers several advantages for our data structure. First, it accommodates the repeated-measures design of our study, which violates the independence assumptions of standard regression models. Second, dengue transmission is influenced by multiple spatially and temporally heterogenous factors, such as land-use, human mobility, and human susceptibility—many of which are unobserved or difficult to quantify and control for directly. Third, the framework accommodates non-linear and lagged effects, enabling realistic representation of delayed and non-linear transmission processes. The Bayesian spatio-temporal framework enables us to capture this unmeasured heterogeneity through latent random effects. Random effects improve model fit while reducing potential bias that could arise when relevant sources of variation are omitted or mis-specified (Lindgren & Rue, 2015; Opitz, 2017; Rue et al., 2009).

Monthly case counts were modeled using a negative binomial distribution to account for overdispersion, with log-population size included as an offset term to account for differences in population sizes among districts. We specified province-month as a first-order cyclic random walk to account for interdependency among months and latent variation associated with regional seasonality. To account for latent spatio-temporal variation, we modeled district-year effects using spatially structured (conditional autoregressive; e.g., unspecified shared environmental features) and unstructured (i.i.d.; district-level heterogeneity not explained by spatial correlation, such as disease management and healthcare differences) random effects, jointly specified as a Besag–York–Mollié (BYM2) model. The i.i.d. random effect is analogous to the random intercept in a mixed-effects model, while the conditional autoregressive (CAR) component represents a spatial random field that encodes neighborhood structure, assuming that adjacent districts are more similar than distant ones. This spatial component leverages the decay of correlation in incidence across space to capture latent spatial autocorrelation not explained by covariates. The model is penalized for unnecessary complexity (using penalized complexity priors): if a fixed effect explains spatial variation, the model shifts variance away from the high-dimensional spatial random effect toward the simpler, two-dimensional fixed-effect relationship. In our specification, we fit a separate spatial random field for each year, thereby allowing spatial dependence structures to evolve over time. As we had gaps in annual data (2012, 2016), year was not included as a temporally autocorrelated term so independent spatial fields were fit for each year.

#### Climate model

The base model was expanded by integrating both established knowledge of dengue dynamics and formal model selection criteria: the Deviance Information Criterion (DIC), the Watanabe-Akaike Information Criterion (WAIC), and the cross-validated logarithmic score (LCPO) (Gelman et al., 2014; Lewis et al., 2014; Spiegelhalter et al., 2014). Guided by this approach, we incorporated key climatic variables known to influence dengue risk globally: elevation (as a proxy for long-term temperature), minimum monthly temperature, long-term drought (annual mean PDSI), and the number of wet days per month (days exceeding the median daily rainfall). As we would expect in a tropical country, elevation and minimum temperature were highly correlated (Spearman’s ρ = -0.85, p < 0.001). To address multicollinearity, we regressed minimum monthly temperature on elevation and used the residuals as our new temperature anomaly variable, representing spatio-temporal deviations in temperature relative to expectations for a given elevation (i.e., long-term climatology) (Supplementary Text 1).

It is well established that temperature has a strong and nonlinear effect on dengue transmission (Athni et al., 2024; Barboza et al., 2023; Caldwell et al., 2021; Childs et al., 2025; Damtew et al., 2023; Gibb et al., 2023; Hidalgo et al., 2026; Lowe et al., 2018; Mordecai et al., 2019).

Accordingly, we included temperature and elevation in all model selection steps, only using a model selection process to first determine the shape and lag structure of their effects. We first evaluated whether elevation had a linear or non-linear association with dengue risk by comparing a simple linear term to a second-order random walk model. Functional form was selected based on information criteria, where a model was preferred if it had at least two of the lowest DIC, WAIC, and LCPO values. We then incorporated residual temperature into the elevation model using a Distributed Lag Non-linear Model (DLNM) to account for delayed, potentially non-linear effects of monthly temperature on dengue risk. DLNMs allow for flexible modeling of both the shape and temporal distribution of climate effects (Gasparrini, 2014; Lowe et al., 2018), and have helped to improve dengue forecasts in Costa Rica (Barboza et al., 2023; Chou-Chen et al., 2023). To identify the appropriate lag structure, we fit eight models with increasing lag windows, ranging from no lag to a lag of up to six months. When lagged predictors were included, models were fit using distributed lag non-linear models; when no temperature lag was included, temperature was tested as either a linear or non-linear effect. We selected the best model using the same information criteria as outlined above.

Water availability is a key determinant of mosquito habitat and has been shown to influence dengue incidence across spatial and temporal scales, though in a complex and context-specific manner (Barboza et al., 2023; Caldwell et al., 2021; Cawiding et al., 2025; Gibb et al., 2023; Harris et al., 2024; Lowe et al., 2018; Nova et al., 2021; Stewart Ibarra et al., 2013). To evaluate its effect, we built on the elevation-temperature model by testing how different combinations of short-term (number of wet days) and long-term (drought; average annual PDSI) water availability metrics improved model fit. In summary, we again tested the effect of monthly precipitation using a DLNM model at increasing monthly windows (0-6 months). For each window, we additionally calculated model fit when annual drought was not included or included as a linear or non-linear term (second-order random walk).

#### Land-cover and land-use drivers

We then tested how land-cover and land-use influenced dengue risk in Costa Rica. First, we included built volume to determine the level of built environment needed to sustain dengue transmission. We additionally tested for land-use/land-cover variables that might point to environments that support dengue transmission outside of cities: specifically, crop cover and forest cover. For each variable, we first individually fit each variable to the climate model using a linear, log-linear, or second-order random walk term. After choosing the function form that best improved model fit, we evaluated model fit for all combinations of land-use variables. We selected the best model using the same information criteria as outlined above.

Importantly, in Costa Rica, agriculture and elevation are highly collinear (Fig. 3; Spearman’s ρ = -0.85, p < 0.001). We used a Z-matrix design to model crop cover, which allowed us to estimate a separate slope of crop cover within each elevation group, addressing issues associated with collinearity and allowing biologically plausible effect heterogeneity across elevation bands (Franco-Villoria et al., 2019; Gómez-Rubio, 2020). We selected three elevation groups by determining the minimum number of bins needed to reduce Spearman’s ρ between elevation and crop cover to < 0.5 within each bin.

Land-cover and land-use effects on vector-borne disease can be highly context-specific (Lambin et al 2010; Tucker Lima et al. 2017), so we evaluated the sensitivity of our flexible, data-driven response (second-order random-walk) to districts with characteristics associated with localized effects. We excluded subsets of districts in turn: (1) low-infrastructure districts, defined as those in the bottom 15th percentile for household access to electricity, sanitation services, and aqueducts based on the 2011 census (INEC, 2011); (2) districts on the Caribbean slope of Costa Rica — specifically Limón Province, Turrialba canton, and Sarapiquí canton — which have distinct climate and socioeconomic conditions relative to central and western-facing regions; and (3) leaving out districts in one province at a time, to test whether results are robust to exclusion of any single geographic region. For each sensitivity model, we assessed changes in the shape and statistical significance of the effect rather than its magnitude, because the magnitude is expressed as a relative risk anchored to the mean dengue incidence of the estimation sample, which shifts when the subset changes.

We evaluated final model fit using Bayesian posterior predictive checks. We evaluated variable multi-collinearity among all variables using Spearman’s ρ (Supp Fig. 11).

#### Variable contribution

We assessed the spatial contribution of each variable using a leave-one-district-out approach, evaluating how omission of each variable affected the model’s ability to predict dengue in an “unseen” district (i.e., a district not used in model fitting). Specifically, we first fit the full model while withholding one district at a time and generated out-of-sample predictions for dengue risk in the withheld district. We then repeated this procedure while omitting each predictor in turn.

The contribution of each variable was quantified as the change in model performance, measured using mean absolute error (MAE) and the relative change in MAE. This framework assesses how each variable contributes to the model’s ability to generalize spatial dengue patterns and explain broad spatial structure. For all out-of-sample predictions, the spatially correlated (CAR) district–year random effect was removed; this step was critical to prevent latent spatial terms from absorbing variation attributable to the focal predictor. Finally, this approach allowed us to map district-specific changes in relative MAE between the ablation and full models, revealing distinct spatial patterns in predictor importance and how well the local variable–dengue relationship conformed to the nationally estimated mean trend.

## Data Availability

Upon publication, all code and aggregated data will by uploaded on GitHub and Dryad.

## Acknowledgements

We thank the Costa Rica Ministry of Health for providing the data underpinning this study. We thank the Costa Rican Center for High Technology (CeNAT) for support for this collaboration. This work was supported by the Stanford Doerr School of Sustainability and its Disease Ecology in a Changing World (DECO) program; University of Costa Rica’s Tropical Diseases Research Center (project C5237); the Stanford Sustainability Accelerator; the Stanford Institute for

Human-Centered Artificial Intelligence (HAI); the Stanford Woods Institute for the Environment; the Stanford King Center on Global Development; the U.S. National Science Foundation (DEB-2011147 with Fogarty International Center; DGE-2146755); and the U.S. National Institutes of Health (R35GM133439).

## Declaration of generative AI and AI-assisted technologies in the manuscript preparation process

During the preparation of this work, the authors used Claude (Anthropic) to assist with manuscript editing and Python and R code development. The authors reviewed and edited the output as needed and take full responsibility for the content of the published article.

