## Supplemental Materials for "Rural dengue dynamics: the interplay of climate, built environment, and agriculture in Costa Rica"

***Supplementary Tables***

**Supplementary Table 1.** Model selection results for temperature variables, including mean elevation and elevation-adjusted monthly minimum temperature. Model selection was conducted to identify (1) the shape of the elevation variable, (2) the shape of minimum temperature in combination with elevation, and (3) the number of lags of monthly temperature that best improved model fit in combination with elevation. RW2 stands for second-order random walk, DLNM stands for distributed lag nonlinear models, and the lag represents the number of lagged months included in the DLNM. DIC = deviance information criterion; WAIC = Widely Applicable Information Criterion; LCPO = Logarithmic Conditional Predictive Ordinate (a cross-validation metric). For each of these model selection criteria, a lower score indicates a better and more parsimonious fit. The best model is indicated in bold font.

| variable | function | temperature lag | DIC | WAIC | LCPO |
|---|---|---|---|---|---|
| base model | | | 147343 | 146366 | 73272.04 |
| elevation | linear | | 146931 | 146135 | 73155.15 |
| elevation | rw2 | | 146970 | 146157 | 73164.63 |
| monthly temperature residual* | linear | 0 | 146579 | 145798 | 72985.48 |
| monthly temperature residual* | rw2 | 0 | 146581 | 145799 | 72986.16 |
| monthly temperature residual* | DLNM | 1 | 145751 | 145364 | 72787.29 |
| monthly temperature residual* | DLNM | 2 | 145546 | 145173 | 72692.26 |
| monthly temperature residual* | DLNM | 3 | 145260 | 144893 | 72550.92 |
| monthly temperature residual* | DLNM | 4 | 145111 | 144747 | 72476.49 |
| monthly temperature residual* | DLNM | 5 | 144988 | 144626 | 72415.75 |
| **monthly temperature residual*** | **DLNM** | **6** | **144914** | **144552** | **72378.55** |

* model also includes elevation modeled as a rw2 function

**Supplementary Table 2.** Model selection results for precipitation variables, including number of wet days in a month (days with precipitation exceeding the monthly median) and annual drought (mean annual PDSI). Model selection evaluated combinations of precipitation and drought variables to determine the optimal functional form and lag structure for monthly wet days. In the distributed lag non-linear model (DLNM), argvar specifies the functional form of the *exposure–response* relationship (e.g., poly1 = first-degree polynomial; poly2 = second-degree polynomial). Arglag specifies the functional form of the lag–response relationship (e.g., ns1 = natural spline with 1 degree of freedom; ns2 = natural spline with 2 degrees of freedom). RW2 indicates a second-order random walk, and the lag value represents the number of lagged months included in the DLNM. Model performance was compared using DIC (Deviance Information Criterion), WAIC (Widely Applicable Information Criterion), and LCPO (Logarithmic Conditional Predictive Ordinate, a cross-validation metric). For each of these model selection criteria, a lower score indicates a better and more parsimonious fit. The best model is indicated in bold font.

| model | precip lag | argvar | arglag | drought | DIC | WAIC | LCPO |
|---|---|---|---|---|---|---|---|
| precipitation rw2 | 0 | | | none | 144758 | 144400 | 72302.91 |
| precipitation rw2 | 0 | | | linear | 144701 | 144397 | 72299.75 |
| precipitation rw2 | 0 | | | rw2 | 144251 | 144126 | 72156.68 |
| precipitation DLNM | 1 | poly2 | ns2 | none | 144839 | 144479 | 72341.3 |
| precipitation DLNM | 1 | poly2 | ns2 | linear | 144783 | 144477 | 72338.42 |
| precipitation DLNM | 1 | poly2 | ns2 | rw2 | 144336 | 144211 | 72198.2 |
| precipitation DLNM | 1 | poly1 | ns2 | none | 144909 | 144548 | 72376.49 |
| precipitation DLNM | 1 | poly1 | ns2 | linear | 144852 | 144547 | 72373.56 |
| precipitation DLNM | 1 | poly1 | ns2 | rw2 | 144406 | 144281 | 72233.64 |
| precipitation DLNM | 1 | poly2 | ns1 | none | 144839 | 144479 | 72341.31 |
| precipitation DLNM | 1 | poly2 | ns1 | linear | 144783 | 144477 | 72338.44 |
| precipitation DLNM | 1 | poly2 | ns1 | rw2 | 144336 | 144211 | 72198.24 |
| precipitation DLNM | 1 | poly1 | ns1 | none | 144909 | 144548 | 72376.5 |
| precipitation DLNM | 1 | poly1 | ns1 | linear | 144852 | 144547 | 72373.56 |
| precipitation DLNM | 1 | poly1 | ns1 | rw2 | 144406 | 144281 | 72233.64 |
| precipitation DLNM | 2 | poly2 | ns2 | none | 144773 | 144410 | 72305.45 |
| precipitation DLNM | 2 | poly2 | ns2 | linear | 144716 | 144408 | 72302.3 |
| precipitation DLNM | 2 | poly2 | ns2 | rw2 | 144267 | 144140 | 72161.17 |
| precipitation DLNM | 2 | poly1 | ns2 | none | 144911 | 144550 | 72377.27 |
| precipitation DLNM | 2 | poly1 | ns2 | linear | 144854 | 144548 | 72374.59 |
| precipitation DLNM | 2 | poly1 | ns2 | rw2 | 144409 | 144284 | 72235.1 |
| precipitation DLNM | 2 | poly2 | ns1 | none | 144773 | 144410 | 72305.48 |
| precipitation DLNM | 2 | poly2 | ns1 | linear | 144716 | 144408 | 72302.3 |
| precipitation DLNM | 2 | poly2 | ns1 | rw2 | 144267 | 144140 | 72161.13 |
| precipitation DLNM | 2 | poly1 | ns1 | none | 144911 | 144550 | 72377.3 |
| precipitation DLNM | 2 | poly1 | ns1 | linear | 144854 | 144548 | 72374.59 |
| precipitation DLNM | 2 | poly1 | ns1 | rw2 | 144409 | 144284 | 72235.04 |

| | | | | | | | |
|---|---|---|---|---|---|---|---|
| precipitation DLNM | 3 | poly2 | ns2 | none | 144656 | 144300 | 72250.81 |
| precipitation DLNM | 3 | poly2 | ns2 | linear | 144601 | 144299 | 72248.27 |
| precipitation DLNM | 3 | poly2 | ns2 | rw2 | 144151 | 144031 | 72106.74 |
| precipitation DLNM | 3 | poly1 | ns2 | none | 144900 | 144540 | 72372.56 |
| precipitation DLNM | 3 | poly1 | ns2 | linear | 144845 | 144540 | 72370.5 |
| precipitation DLNM | 3 | poly1 | ns2 | rw2 | 144402 | 144277 | 72231.85 |
| precipitation DLNM | 3 | poly2 | ns1 | none | 144653 | 144297 | 72249.15 |
| precipitation DLNM | 3 | poly2 | ns1 | linear | 144598 | 144296 | 72246.57 |
| precipitation DLNM | 3 | poly2 | ns1 | rw2 | 144149 | 144028 | 72105.04 |
| precipitation DLNM | 3 | poly1 | ns1 | none | 144898 | 144538 | 72371.54 |
| precipitation DLNM | 3 | poly1 | ns1 | linear | 144843 | 144538 | 72369.52 |
| precipitation DLNM | 3 | poly1 | ns1 | rw2 | 144400 | 144275 | 72230.89 |
| precipitation DLNM | 4 | poly2 | ns2 | none | 144396 | 144044 | 72121.32 |
| precipitation DLNM | 4 | poly2 | ns2 | linear | 144343 | 144044 | 72119.35 |
| precipitation DLNM | 4 | poly2 | ns2 | rw2 | 143897 | 143777 | 71978.76 |
| precipitation DLNM | 4 | poly1 | ns2 | none | 144803 | 144446 | 72325.5 |
| precipitation DLNM | 4 | poly1 | ns2 | linear | 144751 | 144447 | 72324.23 |
| precipitation DLNM | 4 | poly1 | ns2 | rw2 | 144312 | 144188 | 72187.13 |
| precipitation DLNM | 4 | poly2 | ns1 | none | 144395 | 144042 | 72120.32 |
| precipitation DLNM | 4 | poly2 | ns1 | linear | 144341 | 144042 | 72118.36 |
| precipitation DLNM | 4 | poly2 | ns1 | rw2 | 143896 | 143776 | 71977.93 |
| precipitation DLNM | 4 | poly1 | ns1 | none | 144801 | 144444 | 72324.5 |
| precipitation DLNM | 4 | poly1 | ns1 | linear | 144749 | 144445 | 72323.21 |
| precipitation DLNM | 4 | poly1 | ns1 | rw2 | 144310 | 144186 | 72186.11 |
| precipitation DLNM | 5 | poly2 | ns2 | none | 144147 | 143796 | 71996.48 |
| precipitation DLNM | 5 | poly2 | ns2 | linear | 144097 | 143798 | 71995.41 |
| precipitation DLNM | 5 | poly2 | ns2 | rw2 | 143657 | 143536 | 71857.27 |
| precipitation DLNM | 5 | poly1 | ns2 | none | 144658 | 144306 | 72255.4 |
| precipitation DLNM | 5 | poly1 | ns2 | linear | 144610 | 144309 | 72255.05 |
| precipitation DLNM | 5 | poly1 | ns2 | rw2 | 144178 | 144054 | 72120.36 |
| precipitation DLNM | 5 | poly2 | ns1 | none | 144160 | 143810 | 72003.57 |
| precipitation DLNM | 5 | poly2 | ns1 | linear | 144110 | 143812 | 72002.41 |
| precipitation DLNM | 5 | poly2 | ns1 | rw2 | 143671 | 143551 | 71864.41 |
| precipitation DLNM | 5 | poly1 | ns1 | none | 144666 | 144313 | 72259.08 |
| precipitation DLNM | 5 | poly1 | ns1 | linear | 144617 | 144317 | 72258.63 |
| precipitation DLNM | 5 | poly1 | ns1 | rw2 | 144185 | 144061 | 72123.9 |
| precipitation DLNM | 6 | poly2 | ns2 | none | 144065 | 143712 | 71952.67 |
| precipitation DLNM | 6 | poly2 | ns2 | linear | 144017 | 143715 | 71952.51 |
| precipitation DLNM | 6 | poly2 | ns2 | rw2 | 143585 | 143458 | 71817.01 |
| precipitation DLNM | 6 | poly1 | ns2 | none | 144573 | 144218 | 72209.54 |

| | | | | | | | |
|---|---|---|---|---|---|---|---|
| precipitation DLNM | 6 | poly1 | ns2 | linear | 144527 | 144222 | 72209.85 |
| precipitation DLNM | 6 | poly1 | ns2 | rw2 | 144100 | 143970 | 72077.05 |
| precipitation DLNM | 6 | poly2 | ns1 | none | 144137 | 143784 | 71988.66 |
| precipitation DLNM | 6 | poly2 | ns1 | linear | 144089 | 143786 | 71987.99 |
| **precipitation DLNM** | **6** | **poly2** | **ns1** | **rw2** | **143655** | **143528** | **71851.85** |
| precipitation DLNM | 6 | poly1 | ns1 | none | 144615 | 144260 | 72230.37 |
| precipitation DLNM | 6 | poly1 | ns1 | linear | 144568 | 144264 | 72230.5 |
| precipitation DLNM | 6 | poly1 | ns1 | rw2 | 144141 | 144011 | 72097.43 |

**Supplementary Table 3.** Model selection results for shape of land-cover variables (linear and non-linear [log-linear or second-order random walk]) and test of whether the land-cover variable improves the model with only climate variables*. The number of elevation bins to include in the crop cover model was additionally tested. DIC = deviance information criterion; WAIC = Widely Applicable Information Criterion; LCPO = Logarithmic Conditional Predictive Ordinate (a cross-validation metric). For each of these model selection criteria, a lower score indicates a better and more parsimonious fit. The best models for each variable are indicated in bold font.

| variable | function | elevation bins | DIC | WAIC | LCPO |
|---|---|---|---|---|---|
| building volume | linear | | 143598 | 143493 | 71833.75 |
| **building volume** | **log** | | **143504** | **143443** | **71807.94** |
| building volume | rw2 | | 143510 | 143446 | 71809.57 |
| forest cover | linear | | 143640 | 143522 | 71848.33 |
| forest cover | log | | 143649 | 143524 | 71849.95 |
| **forest cover** | **rw2** | | 143626 | 143510 | 71842.55 |
| crop cover | linear | 3 | 143621 | 143486 | 71830.01 |
| crop cover | linear | 5 | 143633 | 143504 | 71839.16 |
| **crop cover** | **log** | **3** | **143552** | **143462** | **71817.56** |
| crop cover | log | 5 | 143577 | 143483 | 71827.93 |

*climate only model = rw2(elevation) + basis temperature residuals (six-month lag) + basis number of wet days (six-month lag) + rw2(annual drought) + province-month random effect + district-year random effect

**Supplementary Table 4.** Model selection results for the combination of land-cover and climate variables* after selecting variable shape. DIC = deviance information criterion; WAIC = Widely Applicable Information Criterion; LCPO = Logarithmic Conditional Predictive Ordinate (a cross-validation metric). For each of these model selection criteria, a lower score indicates a better and more parsimonious fit. The best model is indicated in bold font.

| variables | DIC | WAIC | LCPO |
|---|---|---|---|
| climate | 143655 | 143528 | 71851.81 |
| climate + building volume | 143504 | 143443 | 71807.93 |
| climate + crop | 143566 | 143465 | 71819.03 |
| climate + building volume + crop | 143471 | 143406 | 71788.98 |
| climate + forest | 143625 | 143510 | 71842.34 |
| climate + building volume + forest | 143477 | 143427 | 71799.55 |
| climate + forest + crop | 143525 | 143446 | 71809.58 |
| **climate + building volume + forest + crop** | **143429** | **143394** | **71782.66** |

*climate only model = rw2(elevation) + rw2(temperature residuals) + basis number of wet days (six-month lag) + rw2(annual drought) + province-month random effect + district-year random effect

**Supplementary Table 5.** Final model parameters: median estimate and lower and upper 95% credible interval limits.

| Parameter | Type | Median | CI 0.025 | CI 0.975 |
|---|---|---|---|---|
| intercept | fixed effect | -2.513 | -3.087 | -1.94 |
| elevation | fixed effect | -1.184 | -1.271 | -1.096 |
| temp residual basis v1 l1 | fixed effect | -0.311 | -0.485 | -0.137 |
| temp residual basis v1 l2 | fixed effect | 0.786 | 0.653 | 0.919 |
| temp residual basis v1 l3 | fixed effect | 0.035 | -0.087 | 0.156 |
| temp residual basis v2 l1 | fixed effect | -0.357 | -0.905 | 0.191 |
| temp residual basis v2 l2 | fixed effect | 1.498 | 1.061 | 1.934 |
| temp residual basis v2 l3 | fixed effect | 0.05 | -0.326 | 0.425 |
| temp residual basis v3 l1 | fixed effect | 0.495 | 0.169 | 0.822 |
| temp residual basis v3 l2 | fixed effect | 1.893 | 1.681 | 2.104 |
| temp residual basis v3 l3 | fixed effect | -0.294 | -0.526 | -0.062 |
| precipitation basis v1 l1 | fixed effect | 1.77 | 1.501 | 2.04 |
| precipitation basis v1 l2 | fixed effect | 0.614 | 0.369 | 0.858 |
| precipitation basis v1 l3 | fixed effect | -0.293 | -0.487 | -0.099 |
| precipitation basis v2 l1 | fixed effect | -2.053 | -2.301 | -1.806 |
| precipitation basis v2 l2 | fixed effect | -1.014 | -1.238 | -0.79 |
| precipitation basis v2 l3 | fixed effect | -0.327 | -0.516 | -0.137 |
| log(building volume) | fixed effect | 0.231 | 0.178 | 0.285 |
| Precision Month | hyperparameter | 18.044 | 17.838 | 18.249 |
| Precision District | hyperparameter | 0.403 | 0.400 | 0.406 |
| Phi District | hyperparameter | 0.730 | 0.727 | 0.732 |
| Precision drought | hyperparameter | 0.056 | 0.055 | 0.056 |
| Precision log(crop cover) | hyperparameter | 5.767 | 5.708 | 5.808 |
| Precision log(forest cover) | hyperparameter | 11.850 | 11.769 | 11.967 |

**Supplementary Table 6.** Percentage of districts where inclusion of each focal covariate increased, decreased, or did not change mean absolute error (MAE) by at least 1%. Each model was trained and tested in a leave-one-district-out framework, excluding the CAR component of the BYM2 random effect so that the random effect did not compensate for latent spatial variation otherwise explained by the focal covariate.

| Variable | % improved | % decreased | % unchanged |
|---|---|---|---|
| Drought | 73.40 | 4.10 | 22.50 |
| Precipitation | 55.00 | 15.20 | 29.80 |
| Temperature* | 59.70 | 5.40 | 34.90 |
| Temp residuals | 47.80 | 11.30 | 40.90 |
| Built volume | 46.50 | 10.50 | 43.00 |
| Crop cover | 60.40 | 9.00 | 30.60 |
| Forest cover | | | |

*including both elevation and minimum temperature anomalies

***Supplementary Figures***

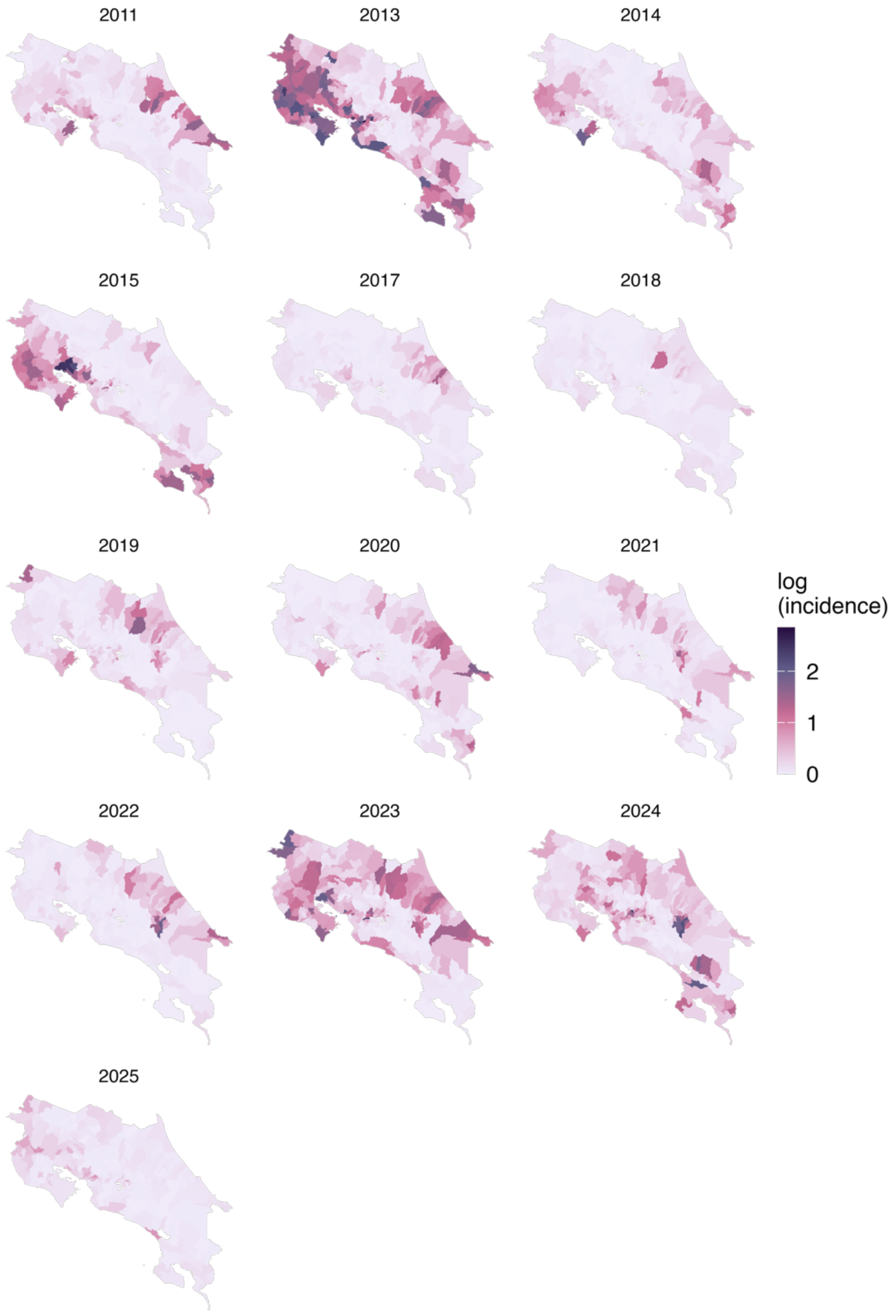


**Supplementary Figure 1.** District-level incidence of dengue per year of the study (cases per 10,000 people). Values are show on the log-scale.

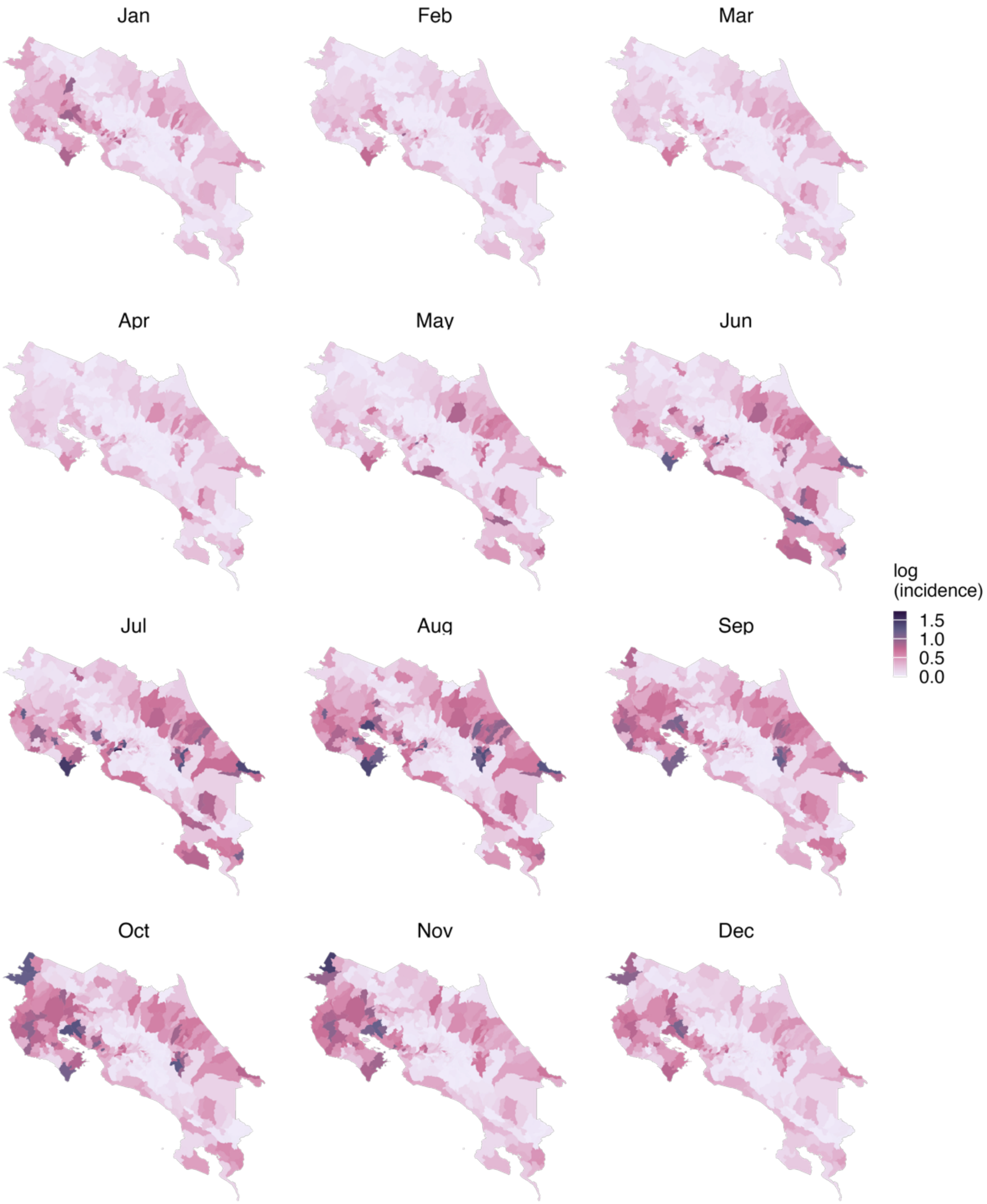


**Supplementary Figure 2.** District-level average incidence of dengue per month over the ten years of the study (cases per 10,000 people). Values are show on the log-scale.

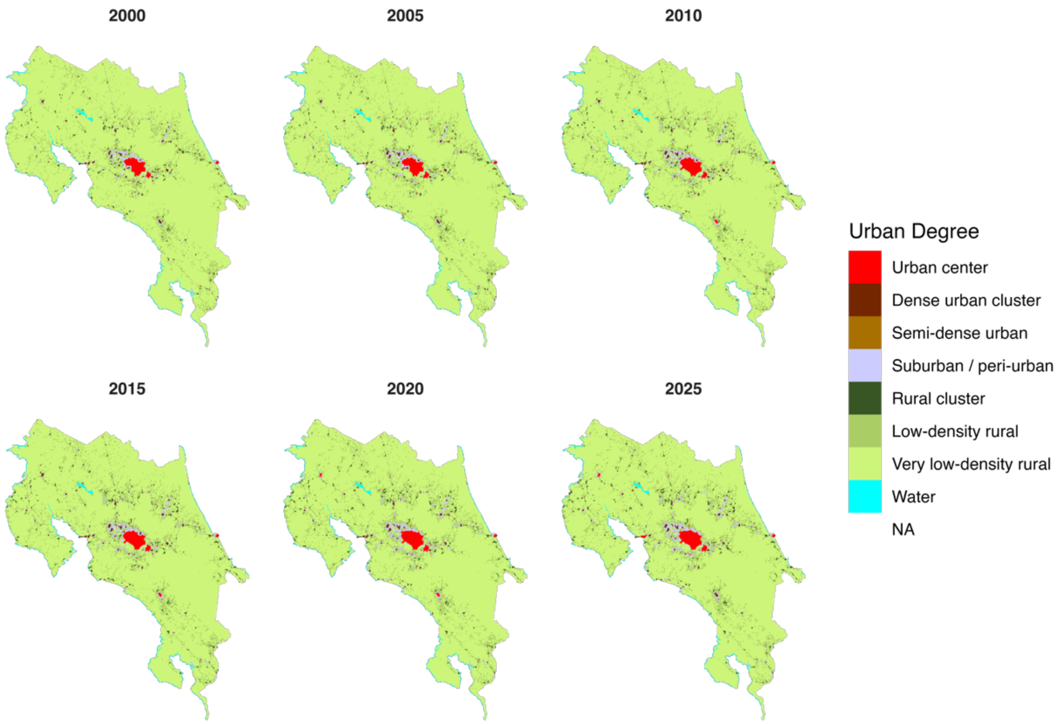


**Supplementary Figure 3.** Urban classification from the Global Human Settlement Layer (Schiavina et al., 2023).

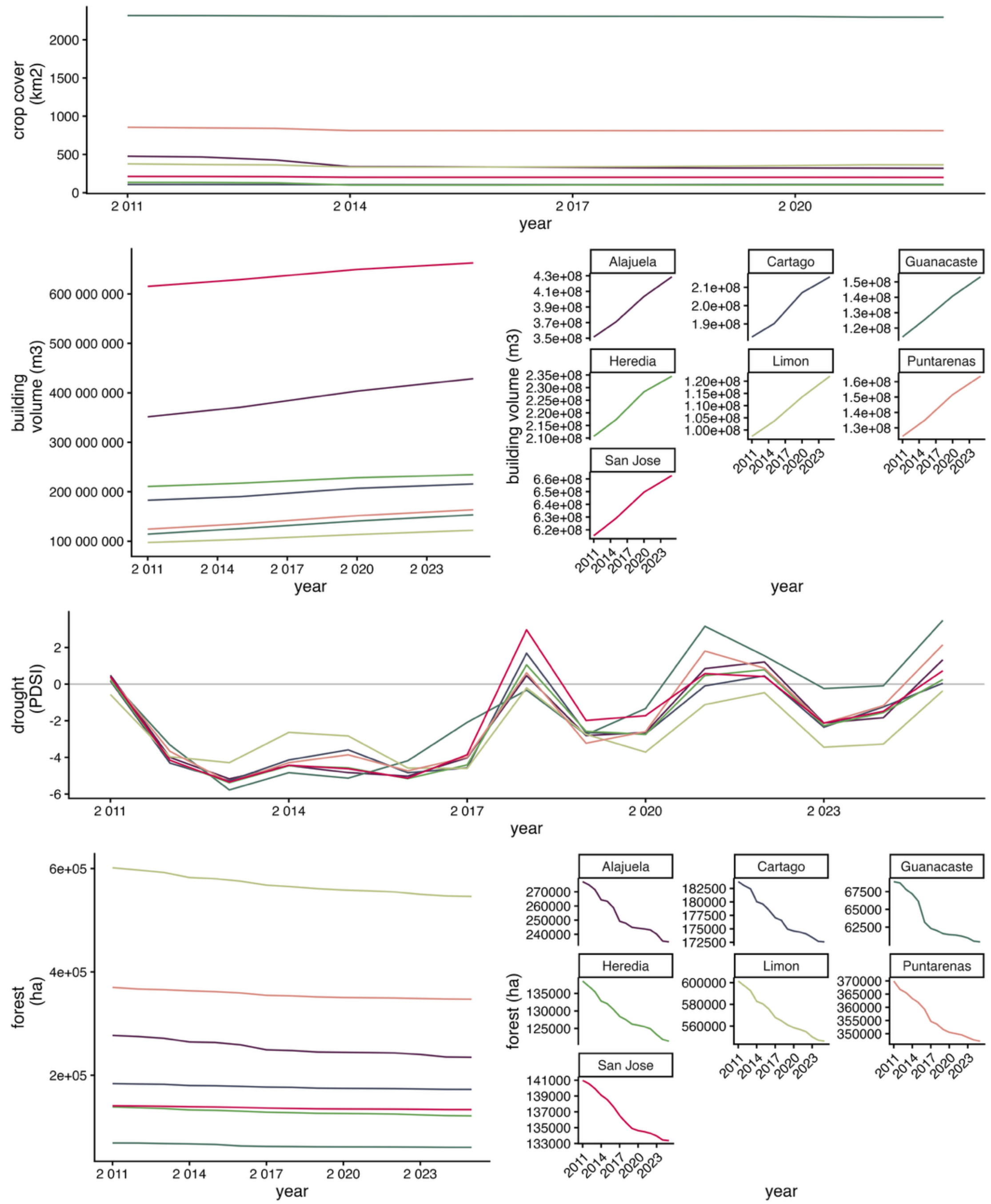


**Supplementary Figure 4.** Average annual crop cover, building volume, and drought per year throughout the study period. Averages are taken over districts. Crop cover is from European Space Agency Climate Change Initiative (CCI) Land Cover data (Defourny, 2019); Building volume from the Global Human Settlement Layer (Pesaresi et al., 2024; Pesaresi & Politis, 2023); PDSI from Terraclimate (Abatzoglou et al., 2018); Forest Cover from JRC Tropical Moist Forest (Vancustsem et al., 2020). Building volume and forest cover shown scaled with provinces relative to each other (left panel) and between the minimum and maximum value for each province (right, faceted panel).

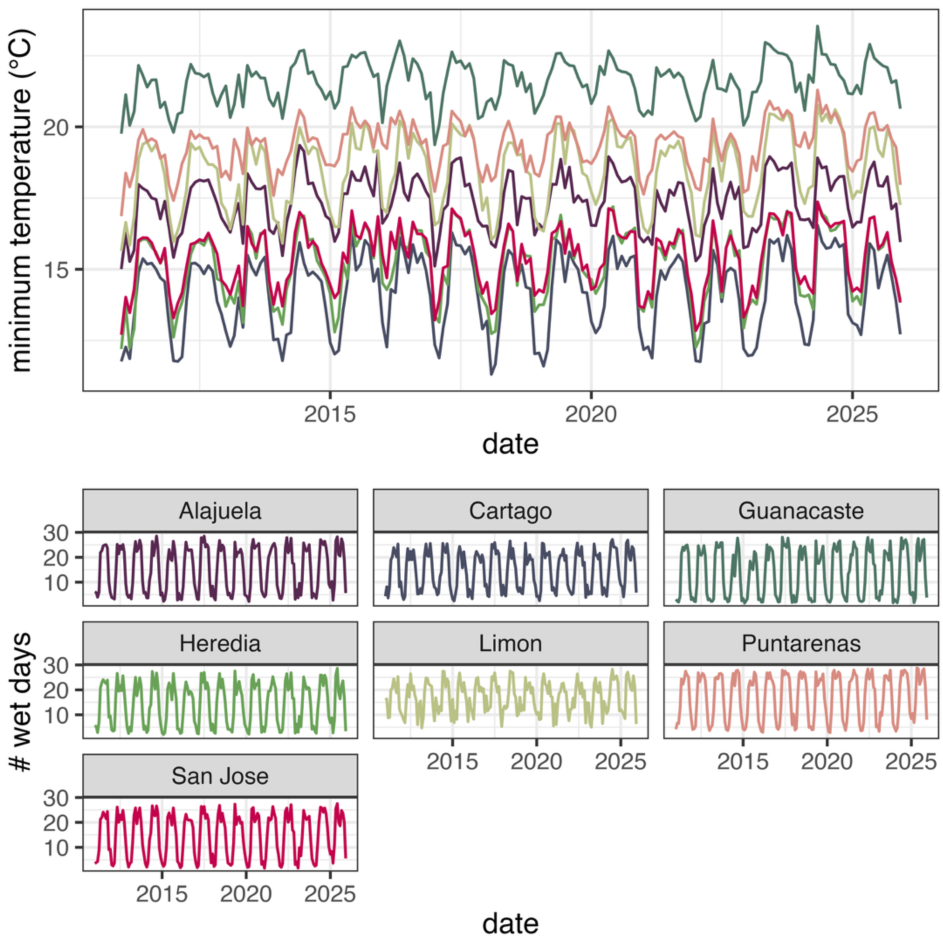


**Supplementary Figure 5.** Average monthly varying covariates (minimum temperature and number of wet days) per month year, average across districts per province. Monthly minimum temperature is from ERA-5 Land (Muñoz Sabater, 2019) and number of wet days is from CHIRPS (Funk et al., 2015).

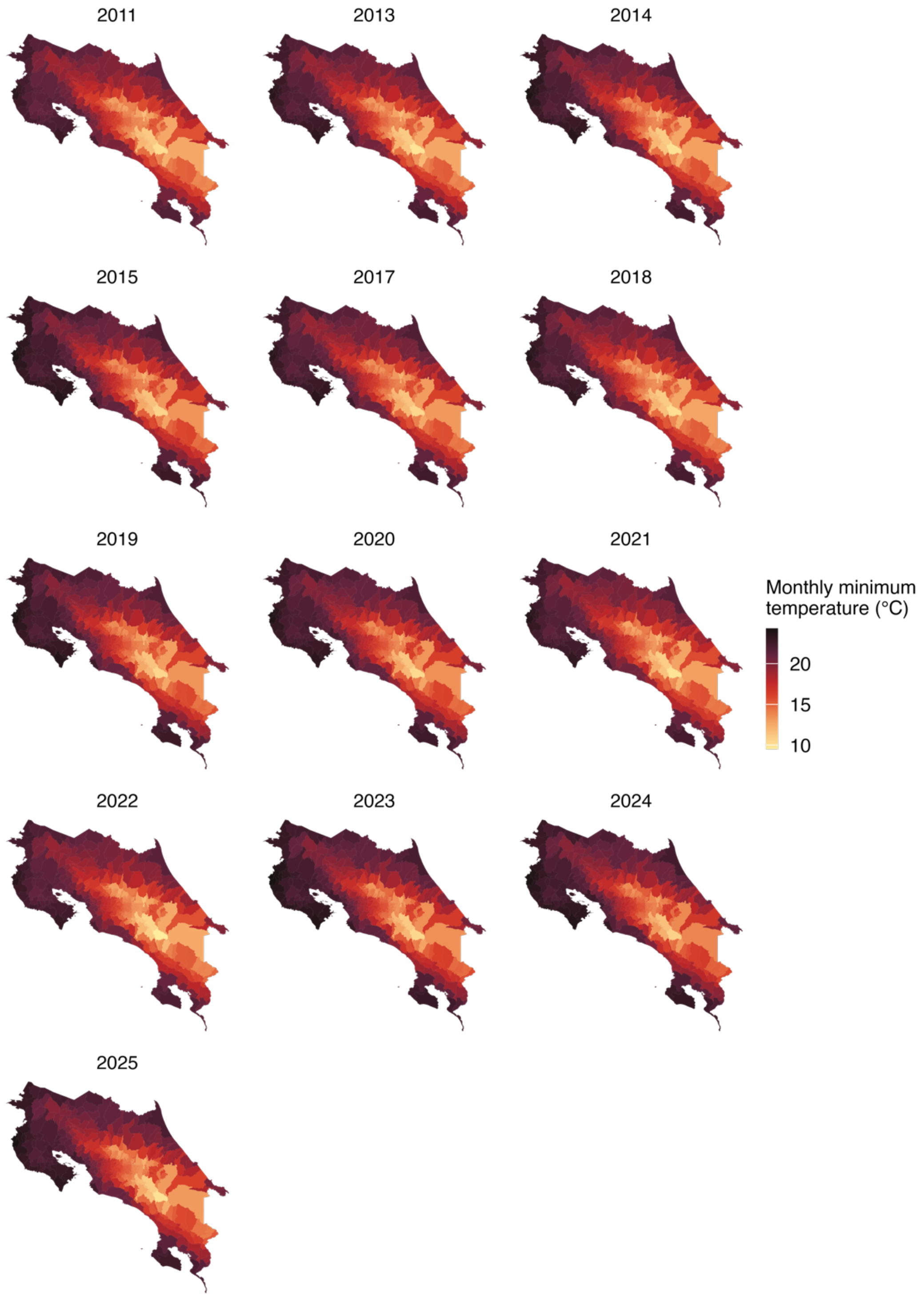


**Supplementary Figure 6.** Average monthly minimum temperature per district-year. Minimum temperature is from ERA-5 Land (Muñoz Sabater, 2019).

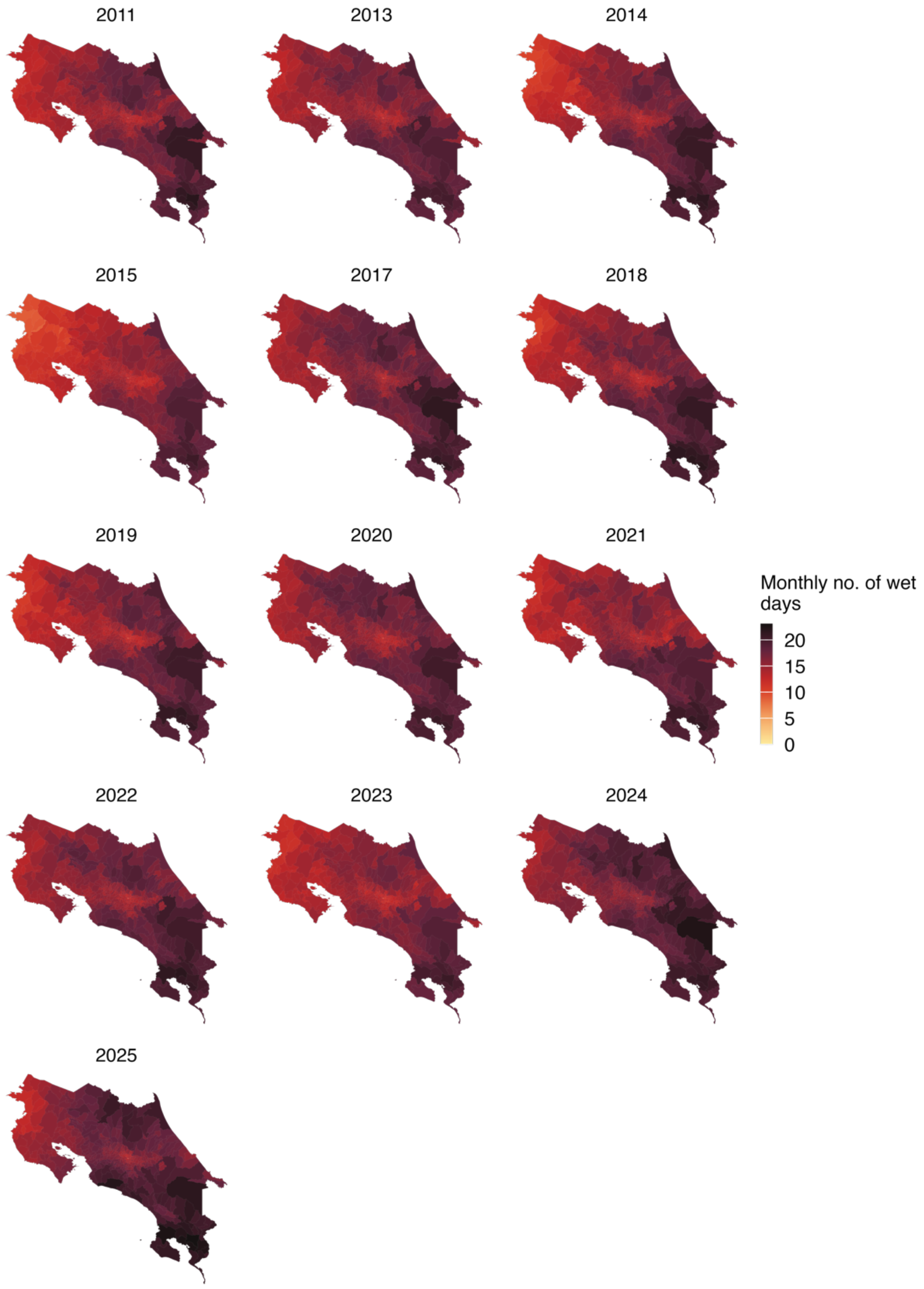


**Supplementary Figure 7.** Average monthly number of wet days (days over the median millimeters of precipitation) per district-year. Number of wet days is based of precipitation data from CHIRPS (Funk et al., 2015).

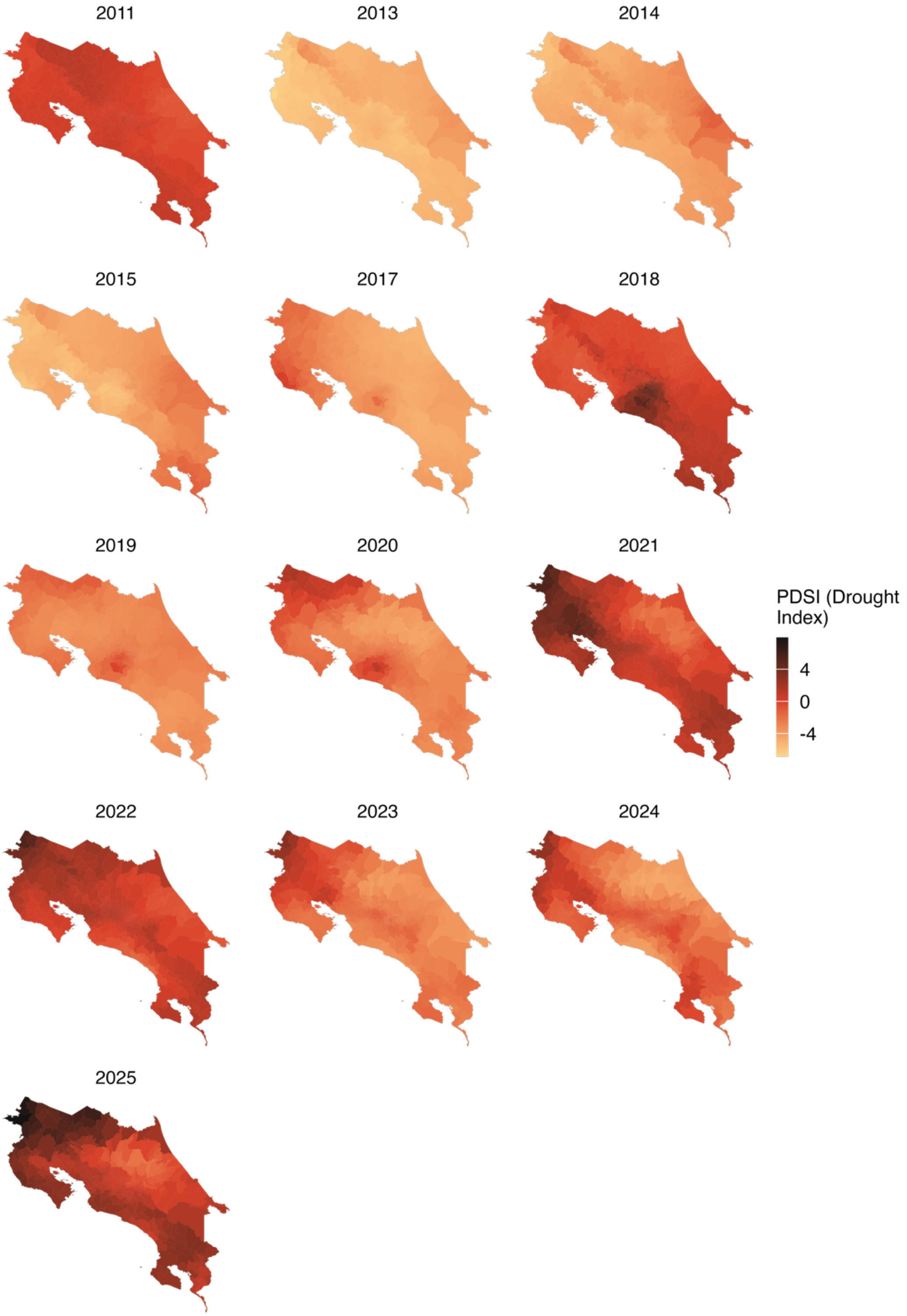


**Supplementary Figure 8.** Annual drought (PDIS) per district-year, where more negative numbers indicate more severe drought. Drought data is from TerraClimate (Abatzoglou et al., 2018).

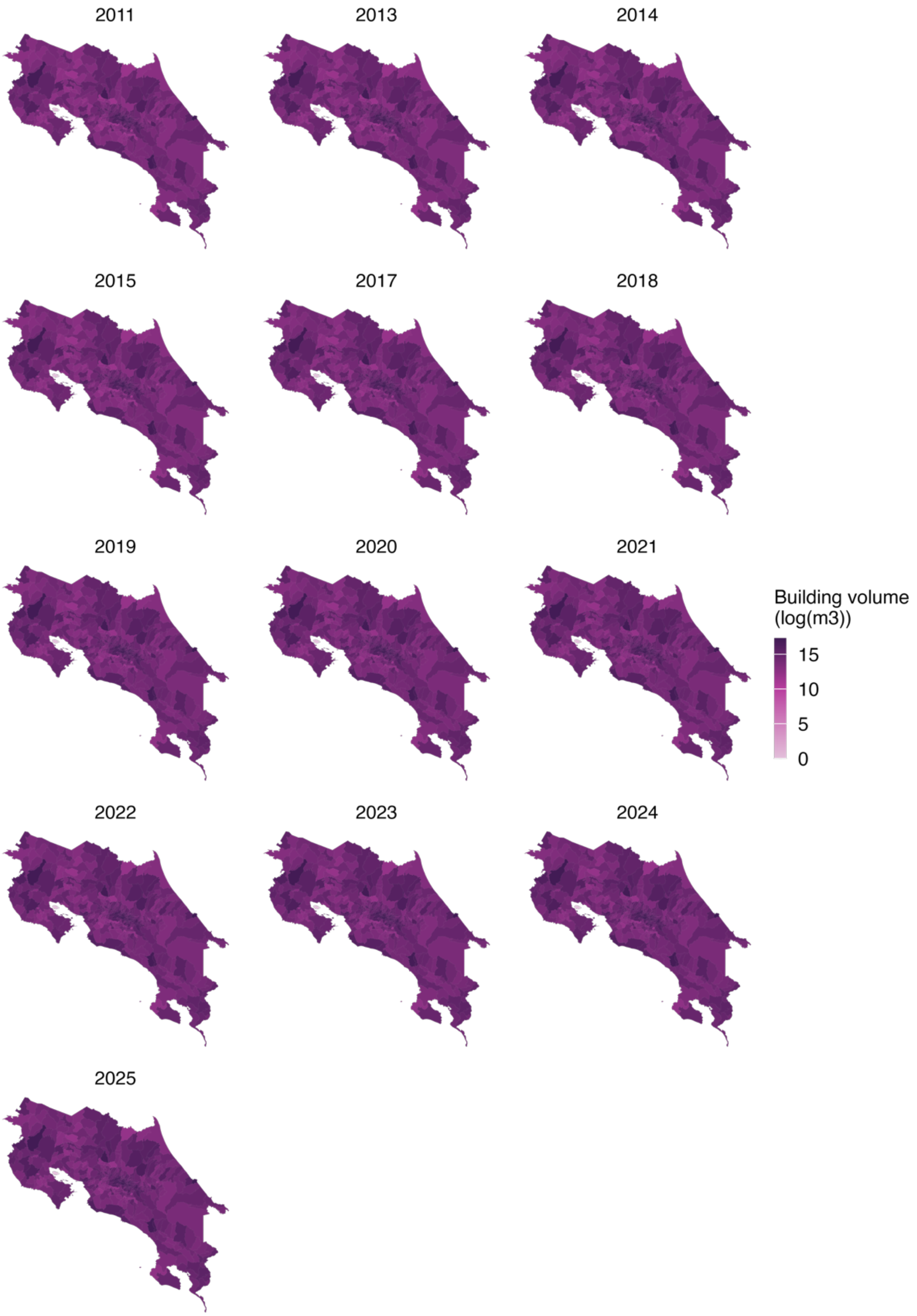


**Supplementary Figure 9.** Building volume ($m^3$) per district-year. Building volume is from the Global Human Settlement Layer (Pesaresi et al., 2024; Pesaresi & Politis, 2023).

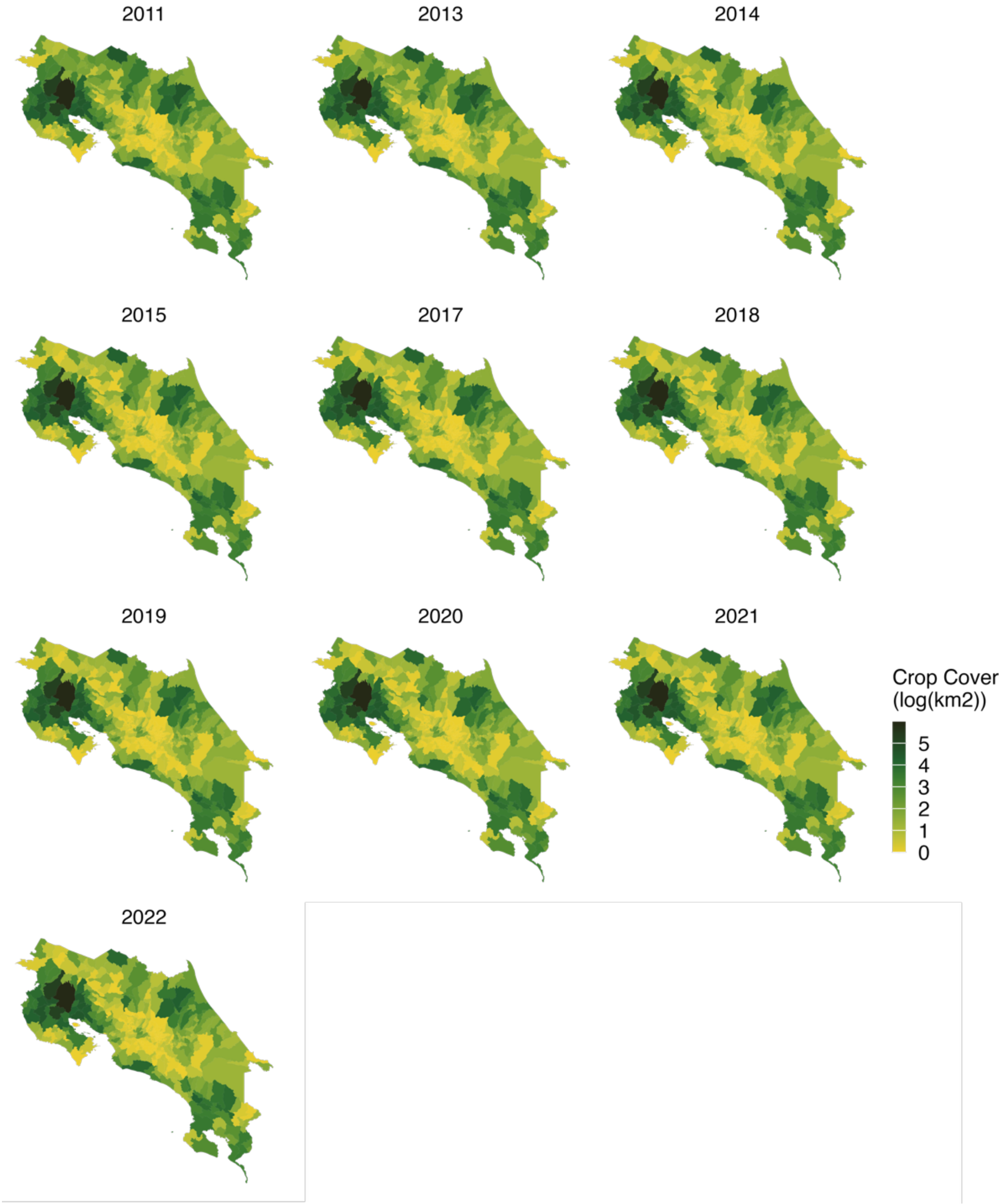


**Supplementary Figure 10.** Crop cover (km$^2$) per district-year. Crop cover is from European Space Agency Climate Change Initiative Land Cover (Defourny, 2019).

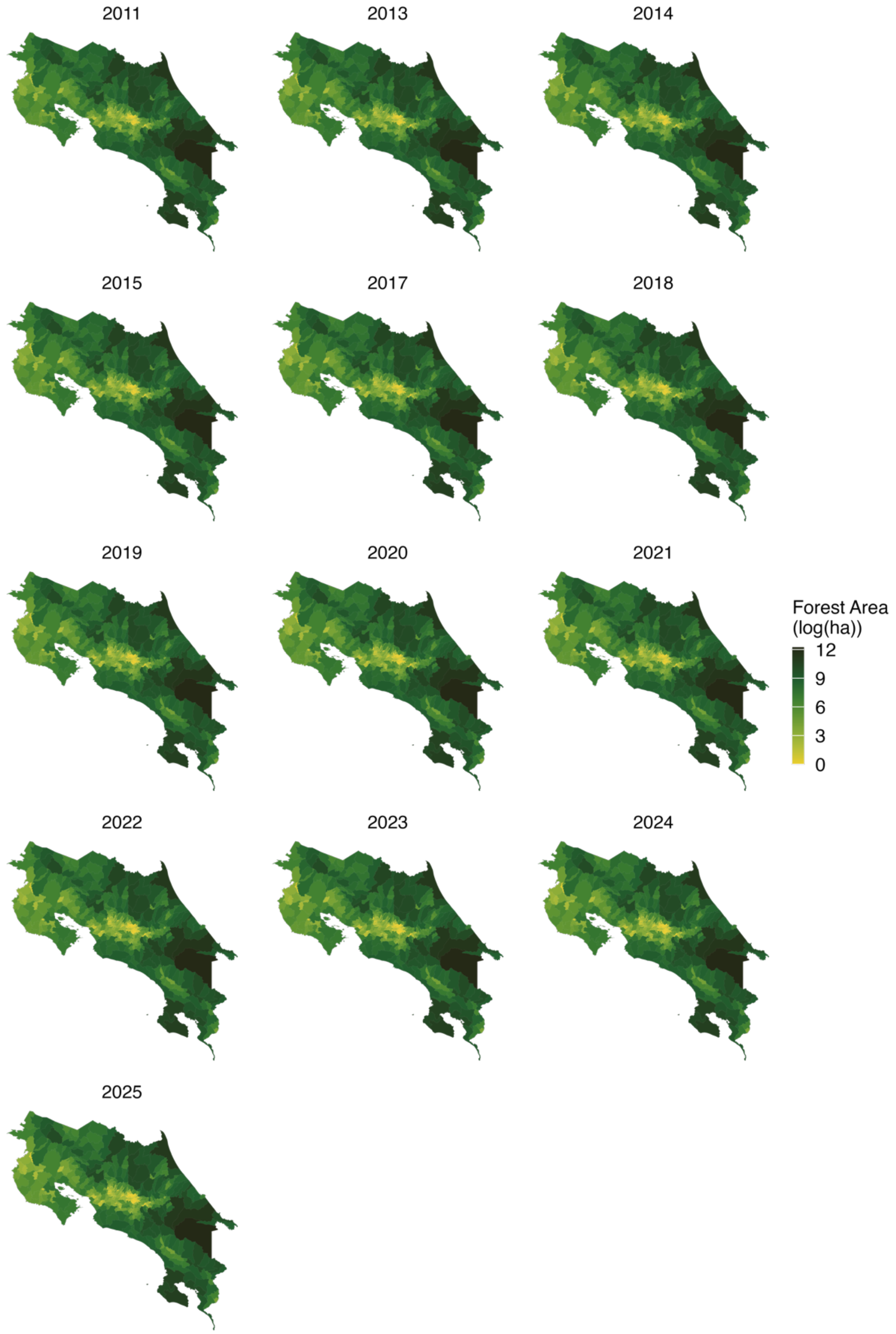


**Supplementary Figure 11. Forest cover (ha) per district-year.** Forest cover is undisturbed forest cover from JRC Tropical Moist Forest dataset (Vancustsem et al., 2020).

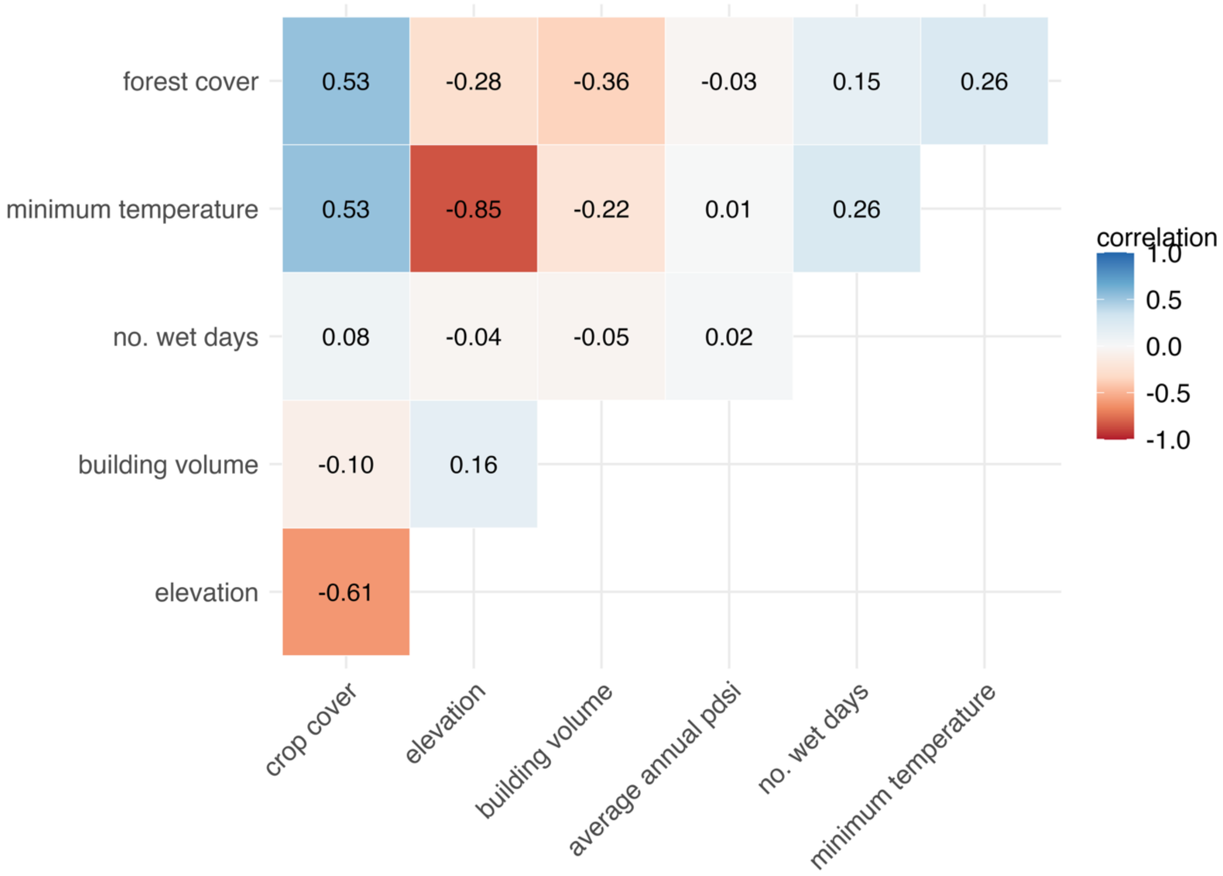


**Supplementary Figure 12.** Spearman's rank correlation between variables.

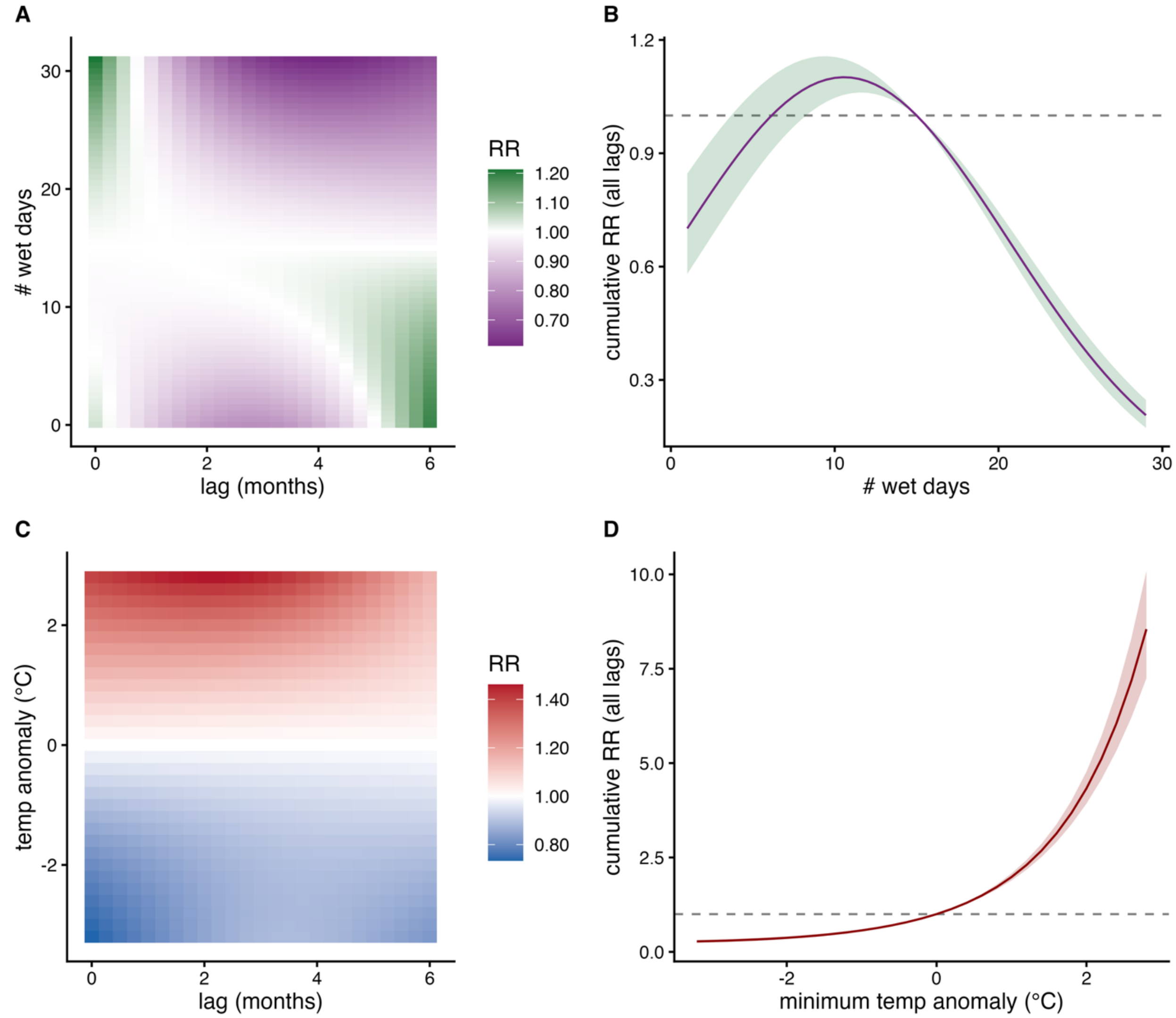


**Supplementary Figure 13. Relative risk (compared to mean value)** per monthly-lag as well as cumulative risk across lags for temperature residuals and precipitation (number of wet days).

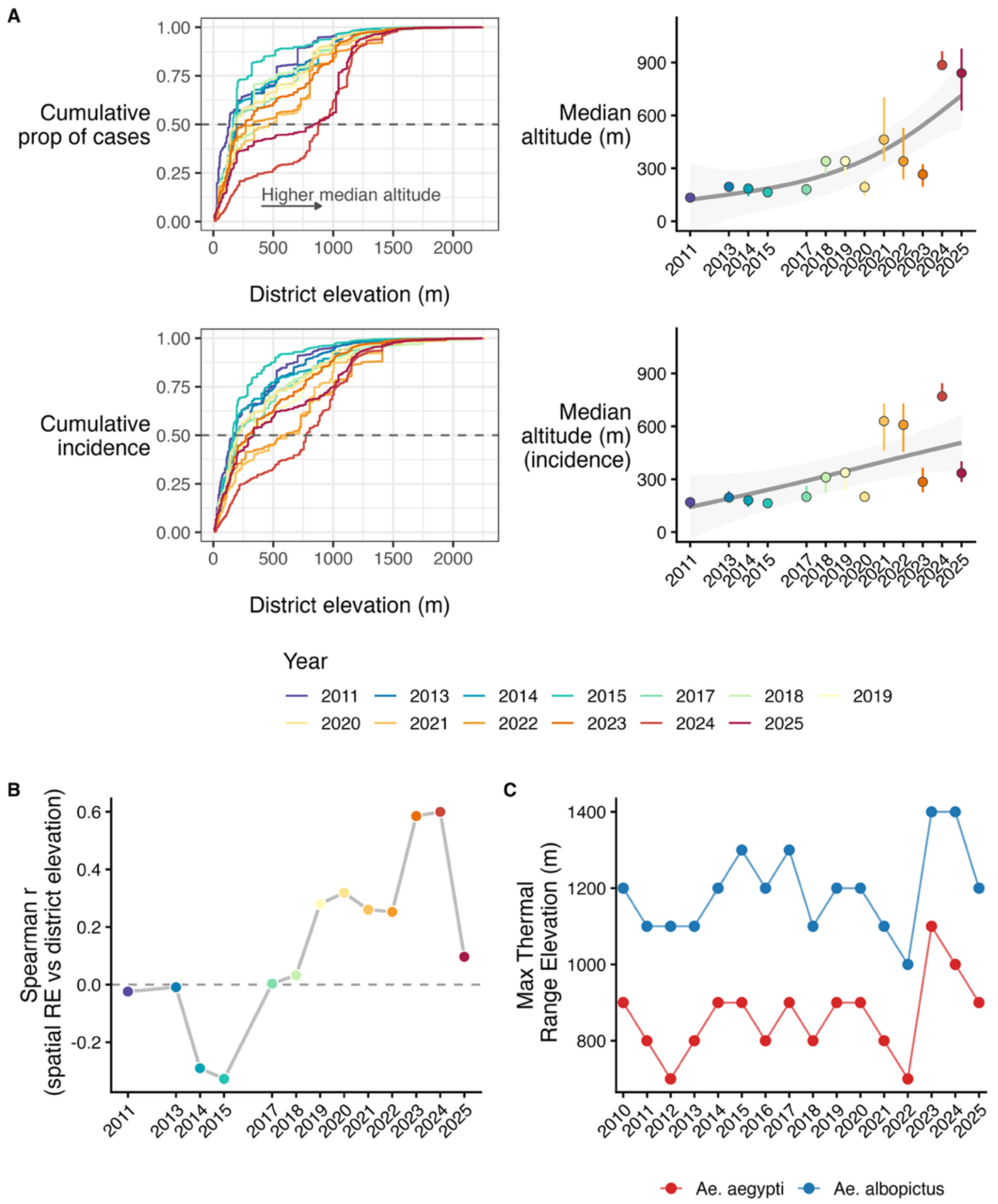


**Supplementary Figure 14. Elevation-associated shifts in dengue burden, 2011–2025.** (A) Cumulative proportion of dengue cases (top) and incidence (bottom) as a function of district elevation for each year (left) and estimated median altitude of dengue burden over time with 95% credible intervals (right), calculated following (Siraj et al., 2014). A rightward shift in cumulative curves and upward trend in median altitude indicate progressive spread into higher-elevation districts. (B) Spearman correlation between district-level spatial random effects from the best-fitting model and mean district elevation, by year. Positive values indicate years in which higher-elevation districts had disproportionately higher incidence. (C) Maximum elevation at which temperature-based relative $R_0$ exceeds zero for *Aedes aegypti* (red) and *Ae. albopictus* (blue) during the June-September transmission season, derived from ERA5-Land minimum monthly temperature (Muñoz Sabater, 2019) and calculated following (Mordecai et al., 2017, 2019).

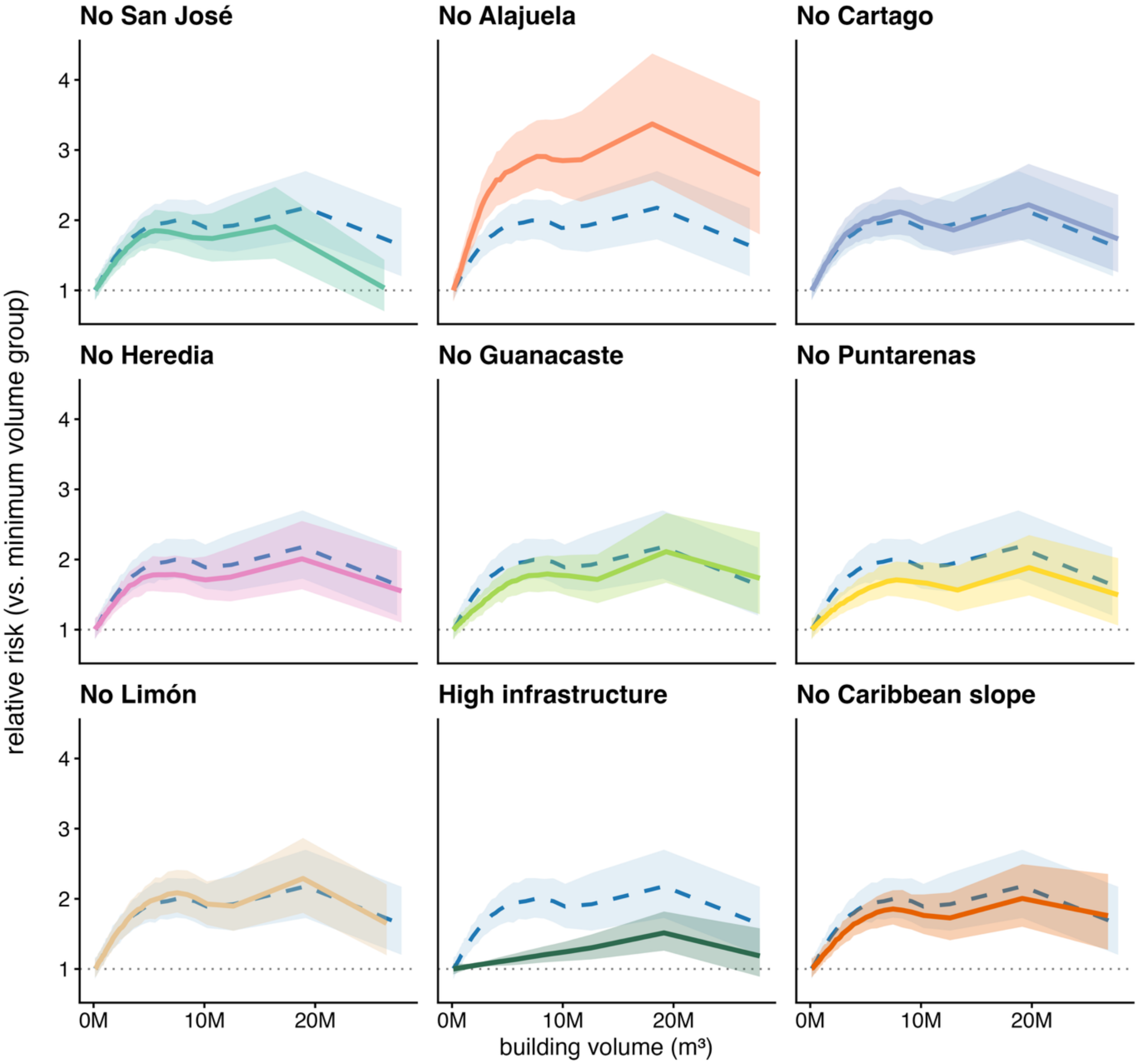


**Supplementary Figure 15. Sensitivity of the estimated building volume–dengue risk relationship to regional exclusion.** Each panel shows the posterior marginal effect of building volume on relative dengue risk after excluding all districts from the named region expressed relative to the median group (reference = 1.0, dotted line). Colored solid lines show the leave-one-out estimate with 95% credible intervals (shaded); the dashed blue line with lighter shading shows the full-data marginal effect for comparison. Panels correspond to Costa Rica's seven provinces (San José, Alajuela, Cartago, Heredia, Guanacaste, Puntarenas, Limón) and two ecological strata (high-infrastructure districts, where low-infrastructure districts were omitted; Caribbean slope districts, including Limón province, Sarapiquí canton and Turrialba canton).

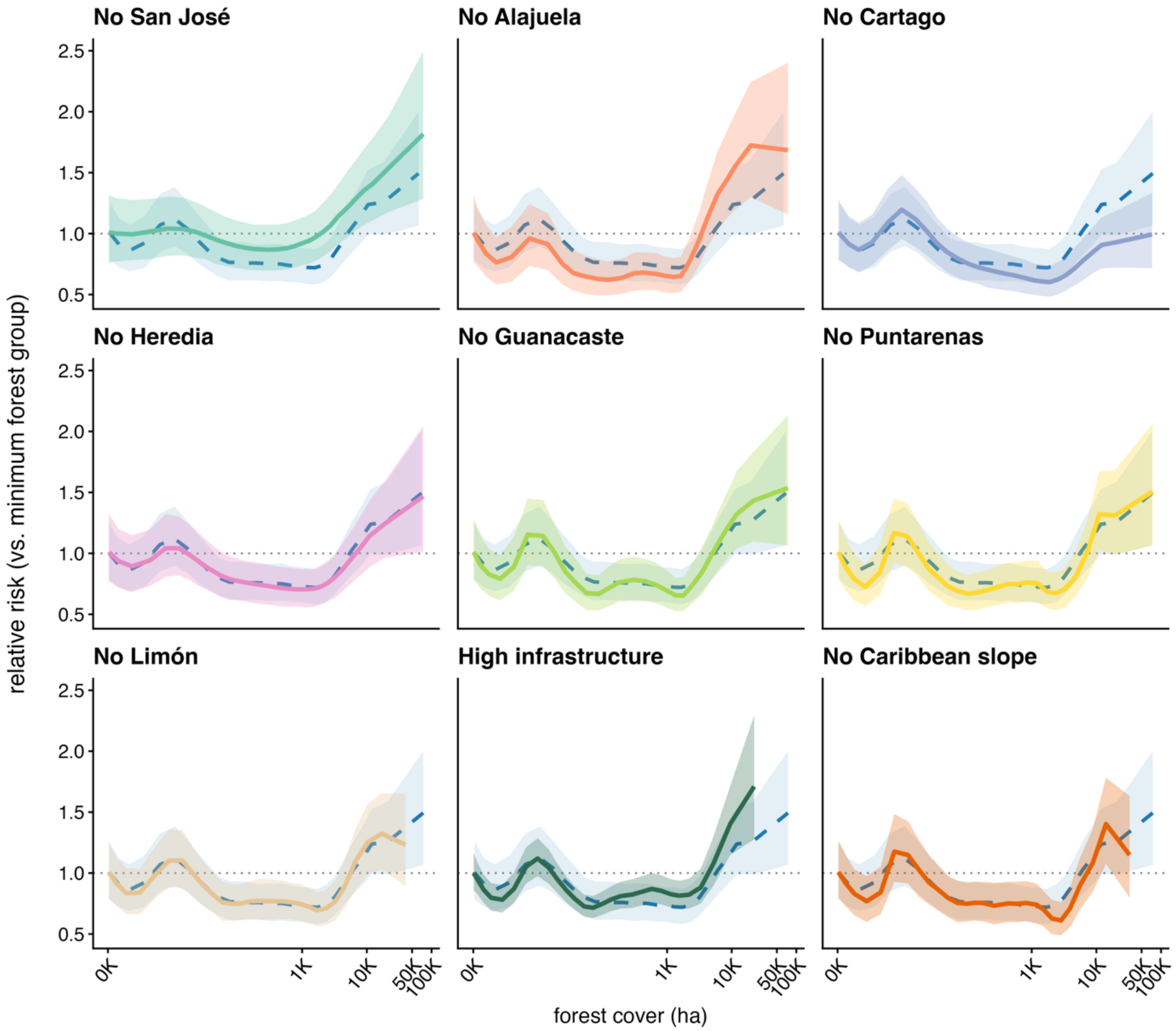


**Supplementary Figure 16. Sensitivity of the estimated forest cover–dengue risk relationship to regional exclusion.** Each panel shows the posterior marginal effect of forest cover (hectares, log scale) on relative dengue risk after excluding all districts from the named region expressed relative to the lowest forest-cover group (reference = 1.0, dotted line). Colored solid lines show the leave-one-out estimate with 95% credible intervals (shaded); the dashed blue line with lighter shading shows the full-data marginal effect for comparison. Panels correspond to Costa Rica's seven provinces (San José, Alajuela, Cartago, Heredia, Guanacaste, Puntarenas, Limón) and two ecological strata (high-infrastructure districts, where low-infrastructure districts were omitted; Caribbean slope districts, including Limón province, Sarapiquí canton and Turrialba canton).

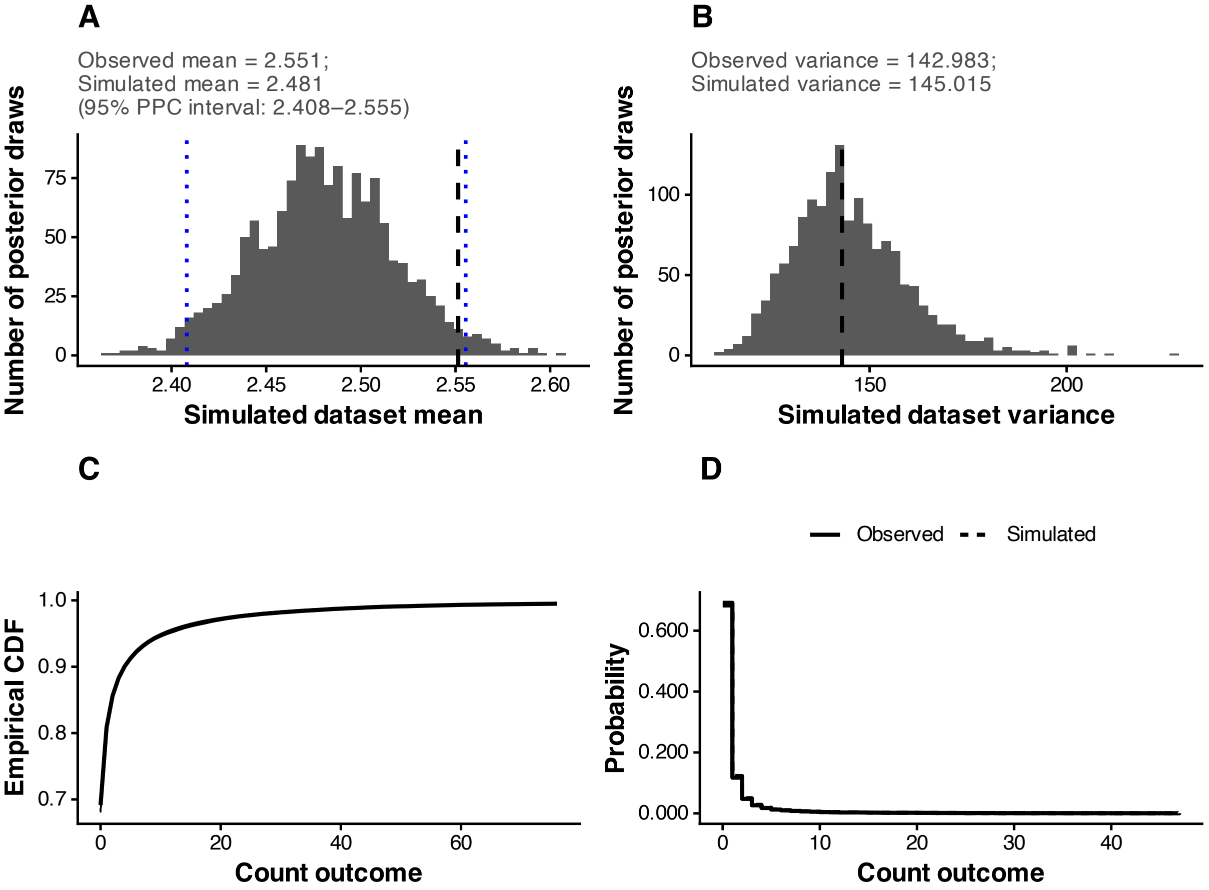


**Supplementary Figure 17. Posterior predictive checks for the dengue incidence model.** Posterior predictive checks were performed by generating 1,000 replicated datasets from the final fitted model and comparing these simulations to the observed data to assess whether the model reproduced key features of the observed distribution. The figure shows the distribution of posterior predictive means across simulated datasets, with the observed mean indicated by the black dashed line and the 95% posterior predictive interval shown in blue (a); distribution of posterior predictive variances across simulated datasets, with the observed variance indicated by the black dashed line (b); Empirical cumulative distribution functions (ECDFs) comparing observed dengue incidence (bold line) with posterior predictive simulations (thin lines) (c); probability mass functions comparing observed and simulated dengue incidence, pooled across posterior draws (d). Panel (a) shows the model predicts dengue counts with a similar, yet slightly lower, mean than the observed data, although this difference was non-significant. Panel (b) shows that the model reproduces the overall variability in dengue incidence observed in the data. Panels (c) and (d) evaluate whether the model produces dengue case counts that look like the real data. The cumulative curves in panel (c) show that the model predicts similar proportions of low, moderate, and high case months as those observed. Panel (d) shows that the model assigns similar probabilities to most case counts, suggesting that it reproduces the overall pattern of dengue incidence, with minor discrepancies primarily for months with no reported cases.

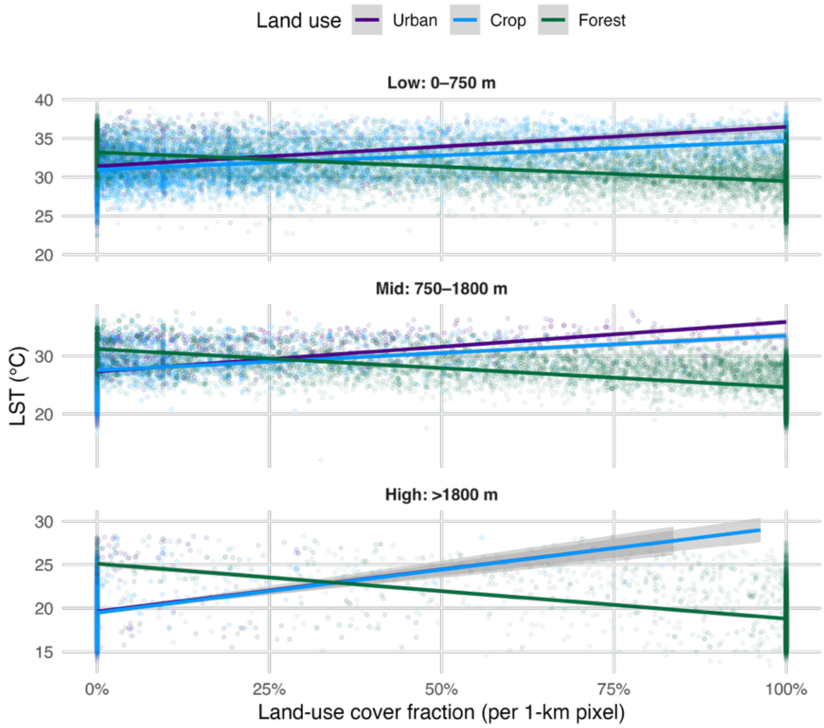


| elevation | Land use | ρ |
|---|---|---|
| Low: 0–750 m | Urban | 0.401 |
| Low: 0–750 m | Crop | 0.443 |
| Low: 0–750 m | Forest | -0.540 |
| Mid: 750–1800 m | Urban | 0.730 |
| Mid: 750–1800 m | Crop | 0.460 |
| Mid: 750–1800 m | Forest | -0.695 |
| High: >1800 m | Urban | 0.285 |
| High: >1800 m | Crop | 0.450 |
| High: >1800 m | Forest | -0.442 |

**Supplementary Figure 18.** Correlation between land surface temperature (LST) and the area of each land-use class across elevation strata. Correlations were calculated using MODIS MOD11A1 Land Surface Temperature (LST) (Wan et al., 2021) and European Space Agency Climate Change Initiative (CCI) Land Cover data (forest, crop, and urban cover) (Defourny, 2019). The native resolution of the land-cover data is 300 m. To match the 1 km resolution of the LST data, we aggregated land-cover data by calculating the proportion of each class within 1 km grid cells. We then randomly sampled pixel values for LST and land-cover fraction across Costa Rica. Sampling was stratified by elevation, land-cover class and by fraction values of > 0.5, ≤ 0.5 and > 0, and = 0 to obtain a balanced distribution across the range of land-use patterns. The figure above shows the relationship between LST and land-cover fraction for each sampled pixel. The accompanying table reports Spearman's rank correlation coefficients between LST and land-cover fraction.

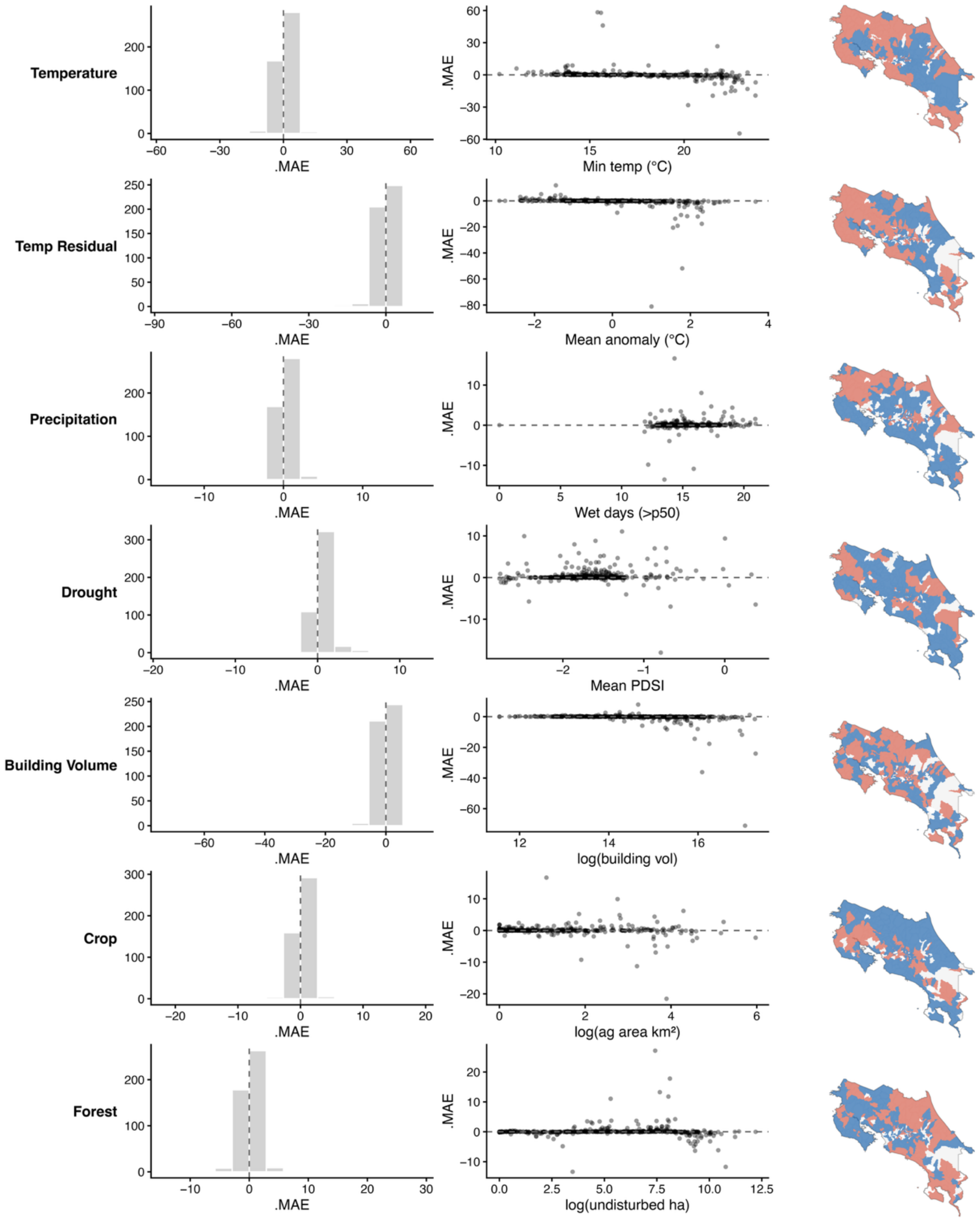


**Supplementary Figure 19.** Change in model error when omitting each focal variable and the latent CAR random effect. Histograms is difference in MAE per district between full model and ablation model. Points are the change in MAE for each district by the average of the variable in the district across the study period. Maps indicate where inclusion of the variable changed relative MAE (($model_{omit\ var}$-$model_{full}$)/$model_{full}$) by at least 1%. We define 'Improved' as relative MAE > 0.01, 'Decreased' as relative MAE < –0.01, and 'No change' as –0.01 ≤ relative MAE ≤ 0.01.

***Supplementary Methods and Results***

Supplementary Text 1. Collinearity between elevation and minimum temperature.

Elevation and minimum temperature were strongly and non-linearly related (Spearman's $\rho = -0.85$, $p < 0.001$). To address multicollinearity, we modeled minimum monthly temperature as a smooth, nonlinear function of elevation and used the residuals from this relationship as a new covariate, representing spatiotemporal deviations in temperature relative to expectations for a given elevation (i.e., departures from long-term climatology). This approach captures variation in temperature across space and time along the elevational gradient while removing the dominant, nonlinear climate–elevation relationship. Using these residuals allowed us to include both long-term climatic structure (elevation) and short-term weather variability (temperature anomalies) in the model without inducing instability from collinearity. In the nonlinear regression of minimum monthly temperature on elevation, elevation alone explained 88% of the variance ($R^2 = 0.88$; Supp. Fig. 16). By including both elevation and the residual temperature term, our model effectively captures absolute thermal conditions for each district and month while partitioning long-term climatological constraints from short-term weather variability.

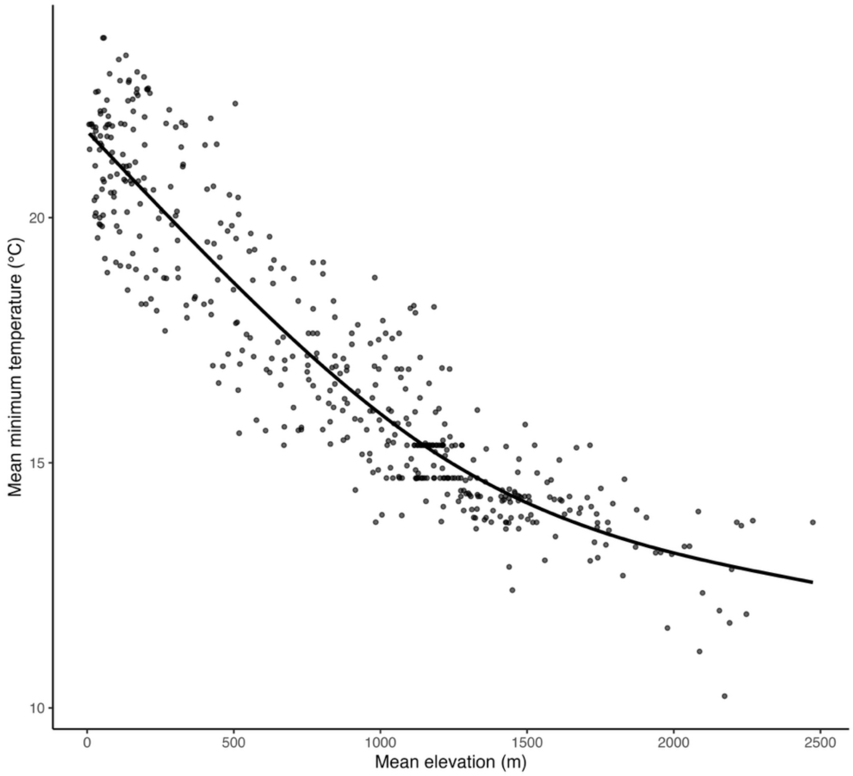


**Supplementary Figure 20.** Relationship between mean elevation and mean minimum temperature across Costa Rican districts. Points show observed district-level mean minimum temperatures, and the solid line shows the fitted nonlinear regression of minimum temperature on elevation used to derive temperature residuals.
